# Temporal Trends in Antibiotic Resistance in Europe, 1998-2019

**DOI:** 10.1101/2023.09.27.23296241

**Authors:** Martin Emons, François Blanquart, Sonja Lehtinen

## Abstract

The emergence of resistant phenotypes following the introduction of new antibiotics is well documented. However, the subsequent dynamics of resistance frequencies over long time periods are less well understood: the extent to which resistance frequencies increase, the rate at which resistance frequencies change, and how this depends on antibiotic consumption remain open questions. Here, we systematically analyse the resistance trajectories emerging from 3,375,774 hospital bacterial isolates sampled from infections in Europe over 1998-2019, representing 887 bug-drug-country combinations. Our analyses support a model in which, after an initial increase, resistance frequencies reach a stable intermediate equilibrium. The plurality (37%) of analysed trajectories were best described as ‘stable’ (neither increasing nor decreasing). The second largest category of trajectories (21%) was those best described as ‘stabilising’ – i.e. showing a transition from increasing frequency to a stable plateau. The antibiotic consumption in a country predicts both the equilibrium frequency of the corresponding resistance and the speed at which this equilibrium is reached. Moreover, we find weak evidence that temporal fluctuations in resistance frequency are driven by temporal fluctuations in hospital antibiotic consumption. Overall, our results indicate that ever increasing antibiotic resistance frequencies are not inevitable and that antibiotic management limits resistance spread. A large fraction of the variability in the speed of increase and the equilibrium level of resistance remains unexplained by antibiotic use, suggesting other factors also drive resistance dynamics.

## 1 Introduction

Antibiotic resistance is a serious public health concern, with an estimated 5 million resistance-associated deaths per year globally [1]. The emergence of resistance phenotypes following the introduction of new antibiotics is well documented [2]. However, once present, the fate of these resistance phenotypes – i.e. the change in the frequency of resistance in the population over longer time frames – is less well understood. Existing studies into temporal trends in resistance frequencies, often focused on specific species-antibiotic (‘bug-drug’) combinations, report a variety of resistance trajectories – including increasing, decreasing, apparently stable, and non-monotonic time courses [3, 4, 5, 6, 7, 8, 9, 10, 11, 12, 13, 14]. A recent study looking at 13 bug-drug combinations across a large number of countries also reported a mixture of increasing and decreasing trends [15].

Beyond characterising overall trends, analysis of temporal trajectories has the potential to provide important additional insights into resistance dynamics. Predictions about medium to long-term resistance trends require an understanding of both the *speed* at which resistance frequencies change and how this speed itself changes – e.g. is there evidence of the rate of resistance increase slowing down as resistance frequencies get higher? In other words, if antibiotic consumption rates remain relatively stable, are resistance frequencies expected to continue rising to eventually reach 100% (‘fixation’) or will we observe stabilisation at intermediate frequencies?

This question of fixation is particularly important as it informs long-term expectations about the burden of resistance and has received considerable attention in the theoretical literature. An intuitive understanding of resistance evolution suggests that fixation should occur if resistance is beneficial enough to emerge in the first place. The simplest models of resistance dynamics predict a logistic rise in resistance frequencies (Figure 1B), at a rate dependent on the population antibiotic consumption and fitness cost associated with resistance [16]. For example, under typical simplifying assumptions [16], if a population is prescribed an average of one course of antibiotics per person per year and if resistance has little cost, the growth advantage of resistance (called ‘selection coefficient’ in evolutionary biology) is of 1 per year. This is enough to shift the frequency of resistance from 1% to 99% in 9 years. A higher cost of resistance leads to a slower increase, but fixation is still expected – provided the cost is not so high that it outweighs the benefit of resistance, in which case resistance would not emerge in the first place. These intuitions also hold for many more complex models of resistance.

**Figure 1.**
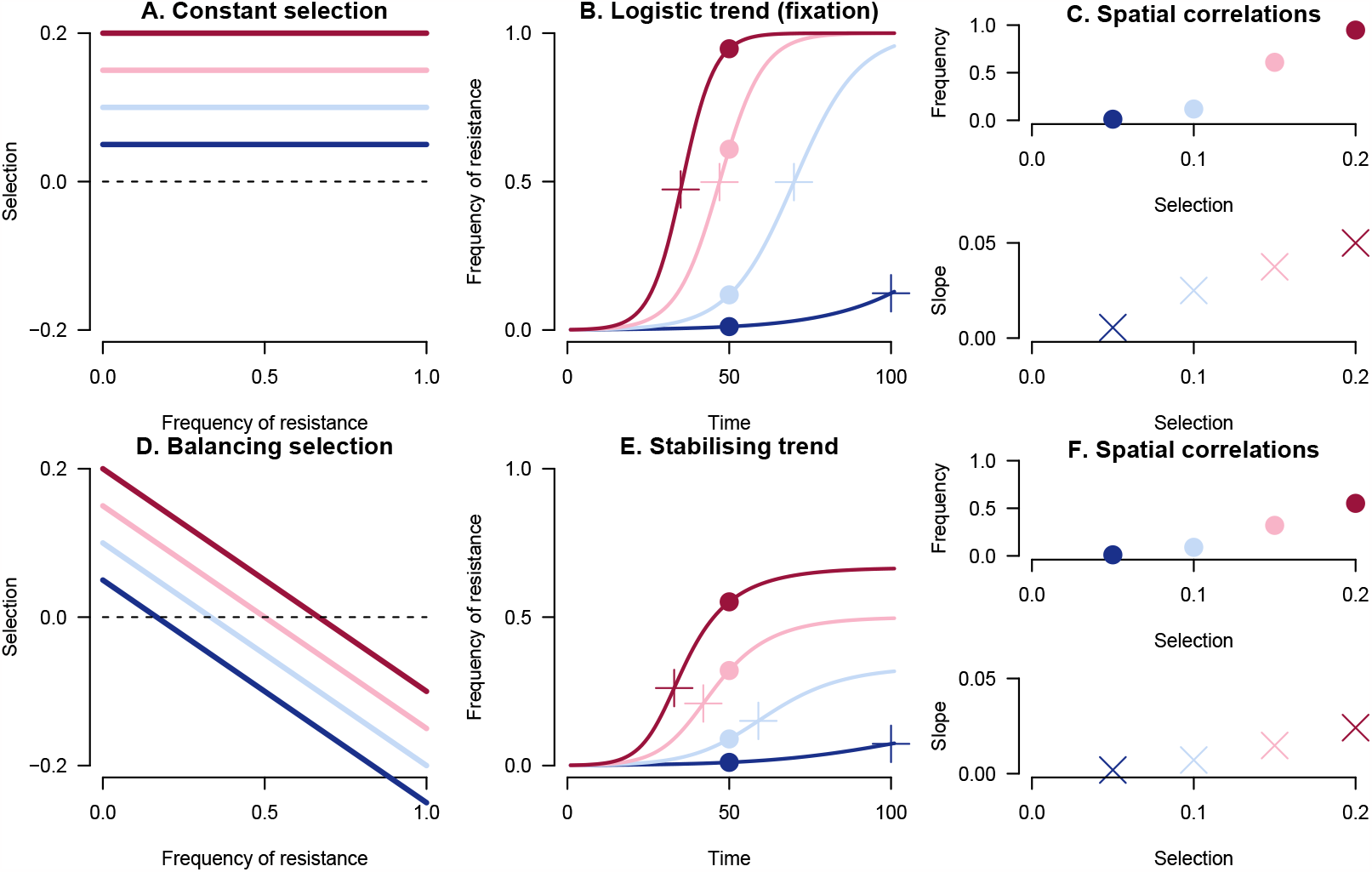
Schematic of constant selection (A, B, C) and balancing selection (D, E, F) models of resistance dynamics. The different colours represent different strengths of selection pressure. For balancing selection, we arbitrarily chose a linearly declining selection coefficient. B and E show the predicted trajectories for each of the models. The frequency dynamics are generally linked with the selection coefficient by the differential equation, *l*?_*p*_ = *s*(*l*_*p*_) where *l*_*p*_ denotes the logit-transformed frequency of resistance (ln[*p/*(1 − *p*)]), with *p* representing the frequency of resistance) and *s*(*l*_*p*_) is the selection coefficient. A simple epidemiological model predicts a constant selection coefficient (A) equal to the antibiotic consumption rate minus the cost of antibiotic resistance [16]. This gives rise to a logistic trend to fixation (B). In contrast, balancing selection (D) leads to a stabilising trend (E). A number of candidate mechanisms lead to balancing selection, but no clear consensus has been reached on a definitive model. C, F: Both models generate a positive correlation between antibiotic consumption and the frequency (here at time 50, materialised by points on B, E) and maximum slope of resistance increase (crosses on B, E).

However, while some resistance trajectories (e.g. penicillin resistance in *Staphylococcus aureus* [4]) have indeed reached fixation, this is not the norm. An alternative model therefore suggests the presence of ‘balancing selection’ (sometimes also called ‘negative frequency-dependent selection’) which acts to stabilise resistance frequencies at intermediate levels. Multiple mechanistic models may generate balancing selection, with stabilising mechanisms arising for example from host population structure [17, 18, 19], strain structure [20] or within-host dynamics [21]. Yet, the lack of observed fixation is not in itself evidence of stabilisation: trajectories could still be rising towards 100%. It is therefore important to systematically assess evidence of stabilization – i.e. a rising phase in resistance followed by a plateau – in temporal trajectories across a range of bug-drug combinations.

Resistance trajectories also matter for understanding the relationship between antibiotic consumption and resistance. Antibiotic consumption correlates with resistance frequencies across European countries and US states [22, 23]. Under the fixation model, this correlation arises because antibiotic consumption affects the *rate* at which resistance increases (Figure 1B,C). On the other hand, under the balancing selection model, the correlation between antibiotic consumption and resistance may arise not only because consumption affects the rate of increase in the rising (‘non-equilibrium’) phase, but also because consumption affects the *equilibrium frequency* of resistance (Figure 1E,F). This distinction matters for predicting the impact of reducing antibiotic consumption. Under the balancing selection model, if resistance is at equilibrium, reduction in antibiotic consumption would lead to a lower plateau. Under non-equilibrium dynamics however, a reduction in antibiotic consumption would either slow or reverse the rise in resistance.

Here we analyse a large number of bacterial isolates sampled from infections in Europe, collected by the European Centre for Disease Control and Prevention (ECDC), together with antibiotic consumption data. We provide a quantitative and systematic view of the temporal trends in resistance and their relation to antibiotic consumption. Firstly, we provide an overview of speed and direction of resistance dynamics and assess evidence for stabilisation. Secondly, we quantify the association between antibiotic consumption and i) the equilibrium frequency of resistance and ii) the speed of increase of resistance. Thirdly, we explore the correlation between year-on-year variation in antibiotic use and resistance. We thus provide a comprehensive picture of antibiotic resistance evolution in Europe over the past two decades.

## 2 Results

### 2.1 Resistance is not systematically increasing and dynamics are slow

We used data from the European Center for Disease Prevention and Control (ECDC) to systematically investigate resistance trajectories in Europe from 1998 to 2019. In brief, the dataset consists of bacterial isolates tested for antibiotic resistance, with data for 8 species (*Streptococcus pneumoniae, Staphylococcus aureus, Enterococcus faecalis, Enterococcus faecium, Escherichia coli, Klebsiella pneumoniae, Pseudomonas aeruginosa*, and *Acinetobacter* spp.), 30 countries and 36 antibiotics. Following data cleaning steps that filtered out trajectories with insufficient sample size, the dataset retained for analysis consisted of 887 bug-drug-country combinations (see Methods for full details).

For an overview of the temporal trends in resistance, we began by fitting a standard logistic model to each trajectory. Overall, increasing trajectories are more common than decreasing ones, with 62% rising vs 38% declining. However, for most trajectories, the temporal trend is not statistically significant at the 0.05 level: 29% of all trajectories are significantly rising and 16% significantly declining (see Figure 2). The median slope – i.e. selection coefficient in the fixation model of resistance – is 0.056 per year for the rising trajectories (-0.051 for the declining trajectories). In the fixation model of resistance, this translates to an increase from 1% to 99% resistance in 165 years.

**Figure 2.**
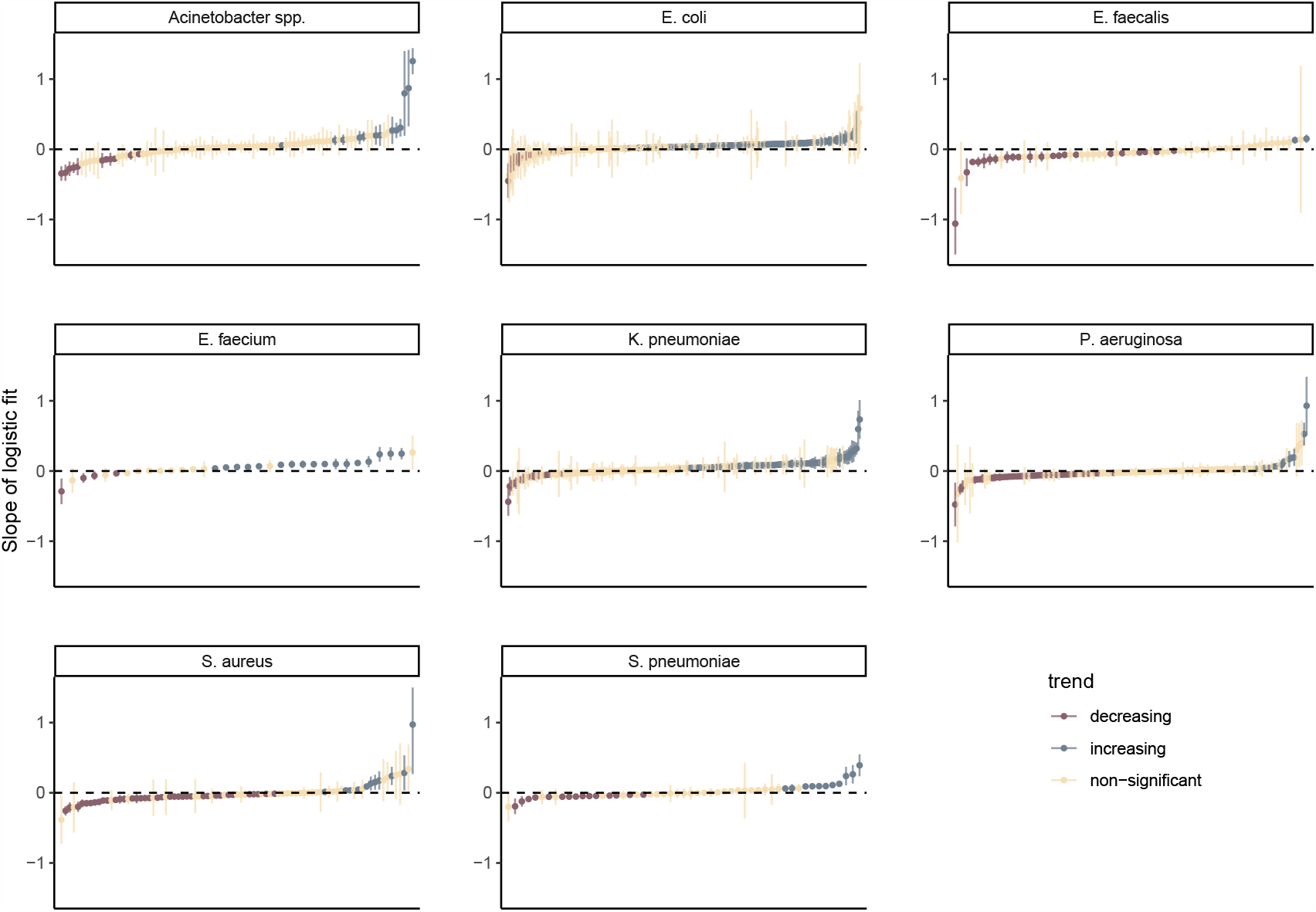
The speed of increase or decrease in European resistance frequencies, as measured by the slope parameter in a standard logistic model, for all bug-drug-country combinations. For clarity, 6 combinations with very high or low slopes (outside of the interval [-1.75;1.75]) have been excluded (see Supporting Information). The plot includes both in- and outpatients. The 95% confidence intervals assume normality.

### 2.2 Evidence of stabilising resistance frequencies

In order to test whether resistance trajectories might be stabilising at an intermediate plateau, we fitted two further models to each trajectory (see Methods for details and SI Figure 1): a scaled logistic model, in which the frequency is increasing towards an intermediate plateau rather than fixation, and a flat straight line with slope 0. Note that 10% of trajectories were not well characterised by any of the models and were removed from further analysis. After exclusion of these, the best model generally fit the data well (Figure 3).

**Figure 3.**
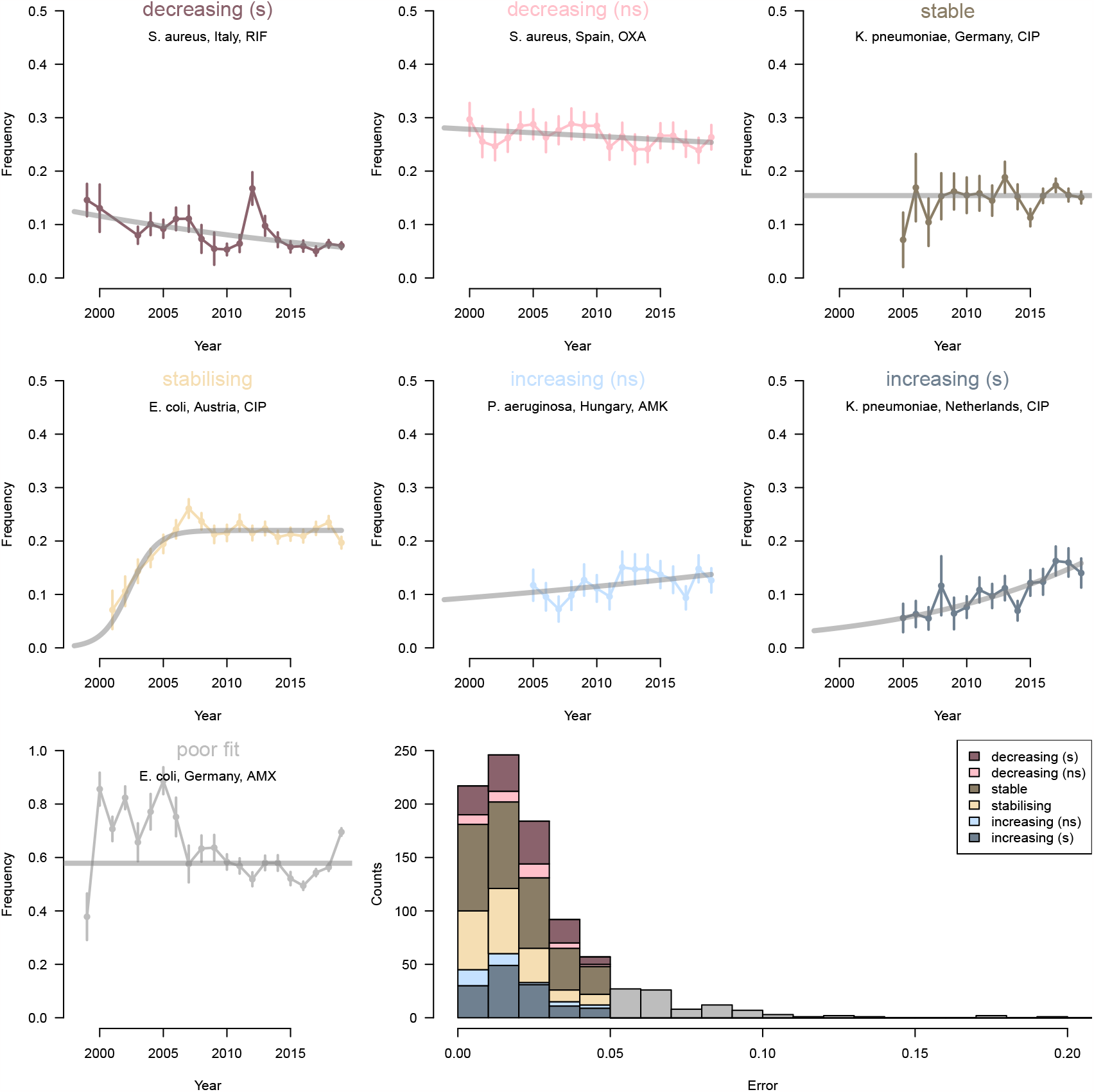
Example trajectories of antibiotic resistance illustrating the different categories. One example (a specific country, bug-drug combination) is shown for each category in the eight panels. The last panels shows the distribution of errors for each category, for all country-bug-drug combinations. Error is quantified as the mean of the absolute value of the deviation between model predicted and actual resistance frequency, and trajectories with poor fit (i.e. error above 0.05) are excluded from further analysis.

We then compared the fits of the standard logistic (2 parameters), the scaled logistic (3 parameters) and flat line (1 parameter) models using the corrected Akaike Information Criterion (AICc). Trajectories best characterised by the plateauing logistic were categorised as ‘stabilising’ (21%) and those best characterised by the flat line as ‘stable’ (37%). The trajectories best characterised by the standard logistic model were further divided into ‘increasing (s)’ (17%), ‘increasing (ns)’ (4%), ‘decreasing (ns)’ (4%) and ‘decreasing (s)’ (17%) categories, where ‘s’ and ‘ns’ denote statistical significance and non-significance at the 0.05 level.

Overall, these results suggest that resistance frequencies have indeed stabilised for many bug-drug combinations (Figure 4): a large proportion of the trajectories which appeared to be increasing in the standard logistic model are now classified as ‘stable’ or ‘stabilising’. This is particularly striking for the species *E. coli* and *K. pneumoniae*. It is worth noting that the stabilising trends give more support to the balancing selection model than the stable trends, because the transition from increasing to stable resistance is observed for these trajectories.

**Figure 4.**
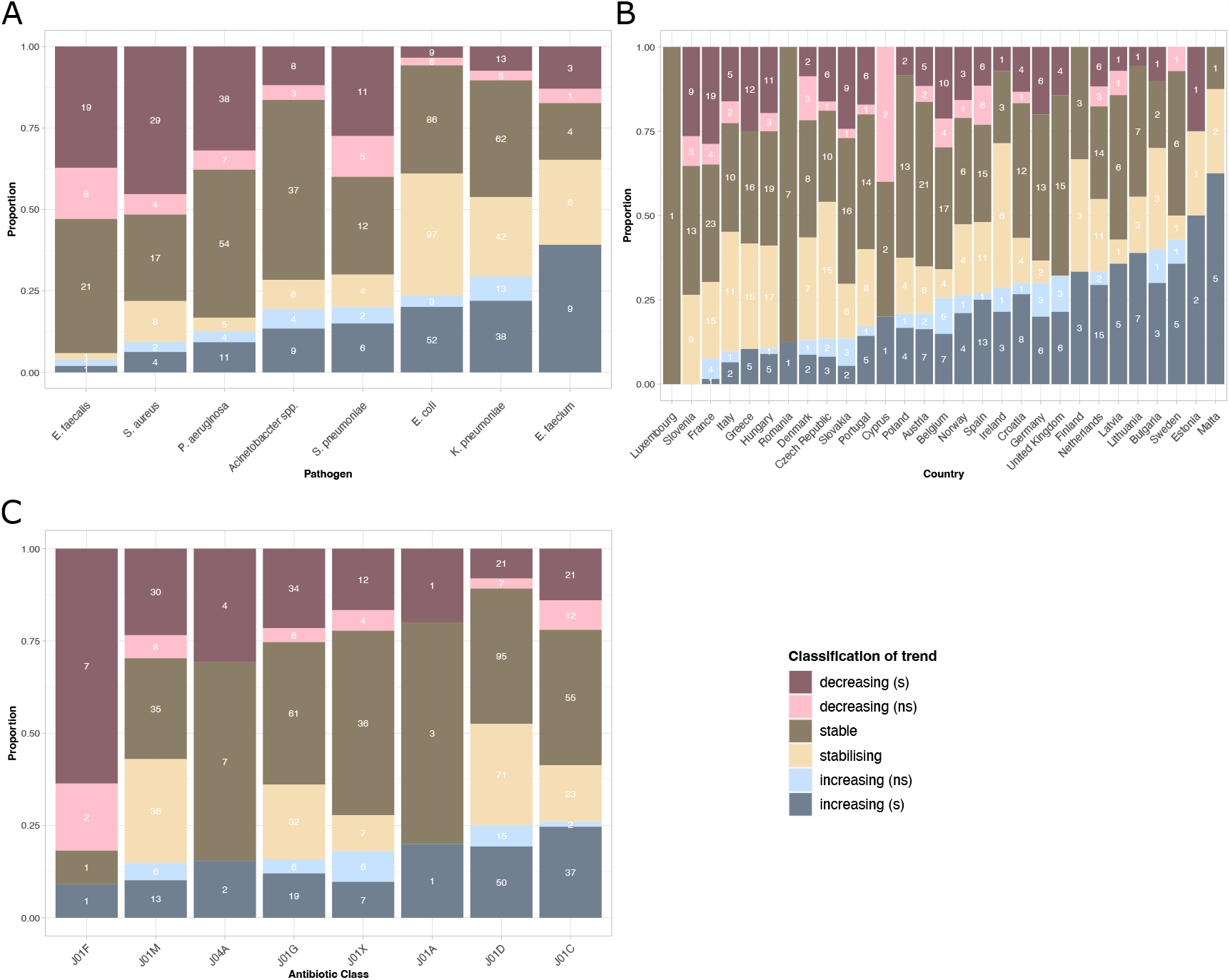
Proportion of trajectories falling into each temporal trend category, by species (A), country (B) and antibiotic class (C). The numbers give the actual sample size of trajectories in each category. This figure does not correct for correlations among the categories (e.g. some species being more frequently sampled in some countries). A corrected analysis is presented in SI Section 2 and is consistent with the information presented here.

To further understand the role of species, country and antibiotic class as determinants of observed temporal trends, we performed a regression analysis (SI Section 2) to correct for correlations among predictors (e.g. some species being more frequently sampled in some countries). The results (SI Figures 2 and 3) are consistent with the information conveyed in Figure 4.

Finally, to gain further insight into the dynamics of the increasing trajectories, we compared the rate at which resistance is rising for the ‘increasing (s)’ category and for the non-equilibrium phase of the ‘stabilising’ category. If the ‘increasing’ trajectories also reflect non-equilibrium dynamics, we would expect these rates to be similar. We quantified the speed of increase for each trajectory as the maximal rate of increase in the time frame for which we had data (Methods 4.4, SI Figure 4). The fastest rates of increase were in the ‘increasing (s)’ category. On the other hand, the mean rate of increase was significantly lower for the ‘increasing (s)’ than the ‘stabilising’ category (2.2 vs 3.9 percentage points per year, *p* = 4.7 *×* 10^−4^ in an linear regression model with ‘stabilising’ vs ‘increasing (s)’ as a predictor of the speed of increase, n = 299). This difference does not arise from confounding, as the effect remains significant in an adjusted model (*p* = 1.9 *×*10^−6^ in a linear regression model with species, country and antibiotic class as additional covariates). Thus while some of the increasing trajectories likely reflect genuine non-equilibrium dynamics, the slow rate of change suggests some of these trajectories may instead represent resistance frequencies responding to a changing selection pressure -due to, for example, increasing antibiotic consumption (see SI Section 3 for caveats).

### 2.3 Consumption of antibiotics predicts the level of stabilisation and the rate of increase of antibiotic resistance

Next, we investigated which properties of the temporal trajectories of antibiotic resistance are predicted by antibiotic use. We correlated both the plateau and maximum rate of increase of the resistance trajectories with the use of the corresponding antibiotic class across countries, for bug-drug combinations for which at least five countries had data (Methods).

First, we observed a positive correlation between the *plateau* level of resistance in the ‘stable’ and ‘stabilising’ trajectories, and the consumption of the corresponding class of antibiotics in the community for most bug-drug combinations (Figure 5A). Overall, the mean correlation was significantly greater than 0 (mean correlation = 0.329 [0.18 − 0.48], *N* = 40 combinations, t-test *p* = 5.2 *×* 10^−5^) and several individual correlations were significantly different from zero, despite considerable uncertainty in each correlation coefficient. This also held true when considering antibiotic use in the hospital instead of in the community (SI Figure 5); mean correlation = 0.322 [0.17− 0.48], *N* = 31 combinations, t-test *p* = 1.7 *×* 10^−4^). This is consistent with selection by antibiotics increasing the plateau level of resistance.

Second, the rate of increase of resistance frequency for the ‘stabilising’ and ‘significantly increasing’ trajectories also exhibited a correlation with antibiotic use, but the signal was weaker than for the plateau. In this analysis, the rate of increase was computed as the maximal slope reached over the time-period in which we had resistance data. The individual correlations could be either negative or positive and none were individually significantly different from 0 (Figure 5B). However, overall, the set of correlations had a mean significantly greater than 0 (mean correlation = − 0.18 [0.06; 0.30], *N* = 14 combinations, t-test *p* = 0.005). The mean was similar when considering antibiotic use in the hospital sector, though not significantly different from 0 (mean correlation = 0.19 [−0.08; 0.45], *N* = 10 combinations, t-test *p* = 0.15) (SI Figure 5). When restricting to stabilising trajectories only, we obtained very few bug-drug combinations, most of them from *E. coli* and showing a positive correlation (SI Figure 6). All in all, there was evidence that the rate of increase in antibiotic resistance is driven by antibiotic use.

**Figure 5.**
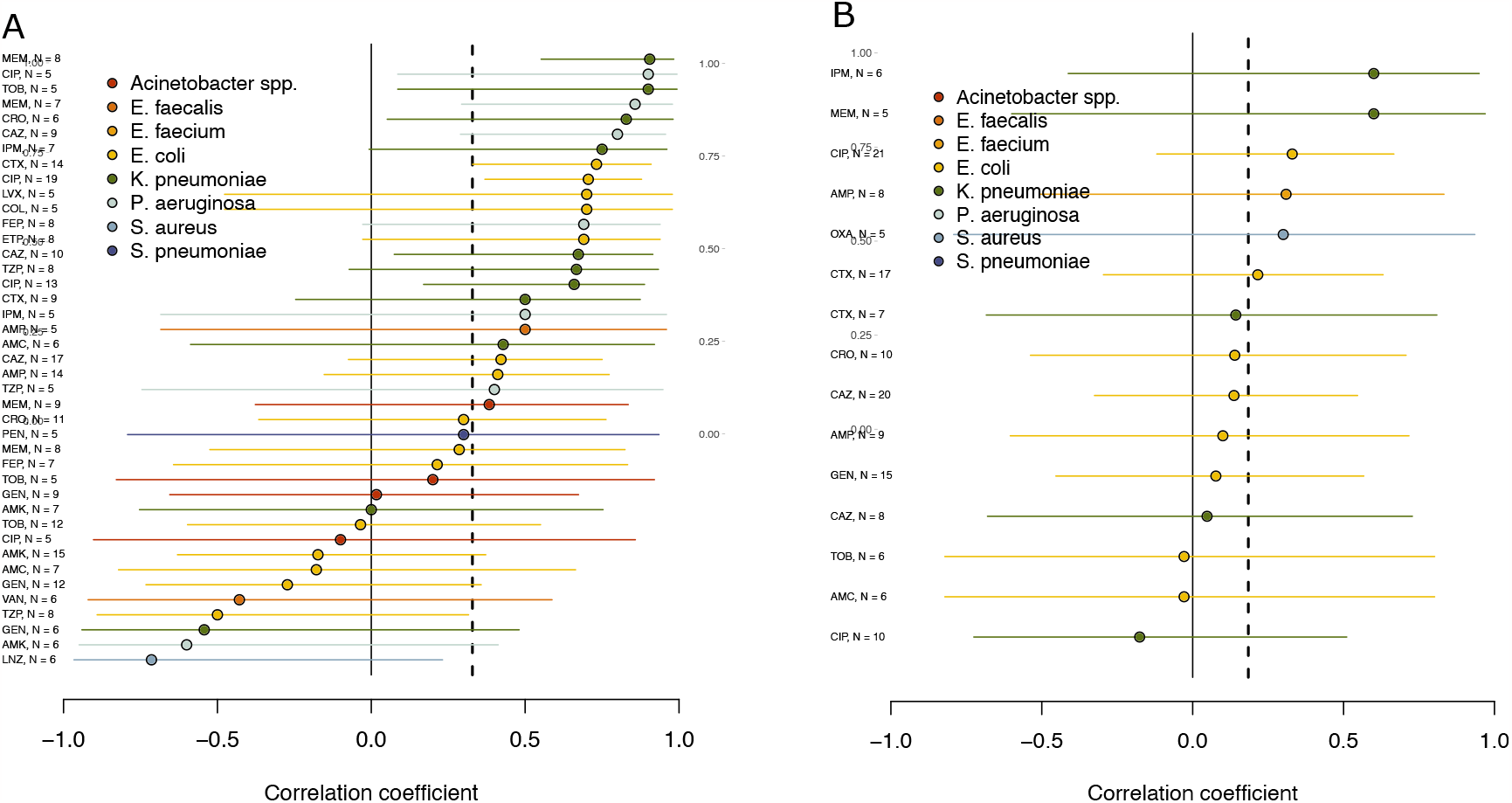
Spatial correlation coefficients between (A) the plateau frequency of antibiotic resistance, or (B) rate of increase, and the rate of use of the corresponding antibiotic in the community, for all bug-drug combinations. The number of countries included is indicated for each combination. The vertical dashed lines show the overall mean. Trends are similar when considering hospital instead of community use (SI Figure 5).

**Figure 6.**
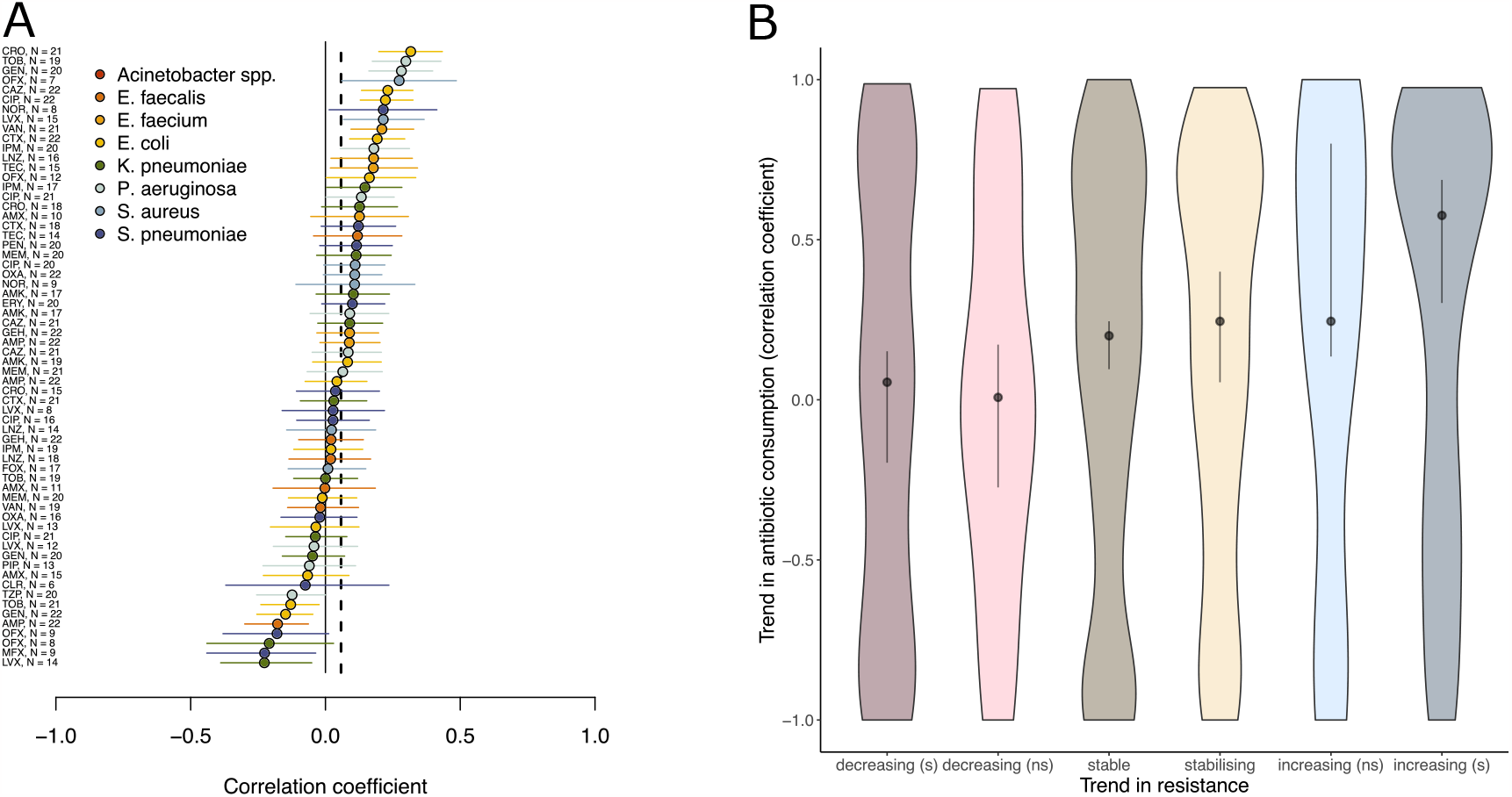
Temporal variation in hospital antibiotic use as a predictor of temporal variation in antibiotic resistance. (A), the temporal correlation of the frequency of resistance with hospital antibiotic use across bug-drug combinations. The number of countries included is indicated for each combination. The vertical dashed lines show the overall mean. (B), the distribution of temporal trends in antibiotic consumption in hospitals for each category of trend in resistance.

Lastly, if we ignore temporal trends, the median level of resistance over time was also generally positively correlated with consumption (SI Figure 7).

### 2.4 Antibiotic use as a predictor of temporal variation in antibiotic resistance

Following the earlier result that ‘increasing’ trajectories may reflect a changing equilibrium frequency, we investigated whether year-to-year variation in antibiotic use could cause corresponding year-to-year fluctuations in the level of resistance. For each country, we computed the temporal correlation of resistance with use of the corresponding antibiotic, for combinations for which we had at least five years of data and for all categories of temporal trajectory. We then averaged these temporal correlations over all countries and retained only bug-drug combinations for which we had at least five countries. There was overall a positive mean correlation between resistance and antibiotic use in the hospital sector (mean correlation = 0.06 [0.03; 0.09], *N* = 63 combinations, t-test *p* = 6.8*×* 10^−4^) (Figure 6A), but this was not true when considering the use of antibiotics in the community sector (mean correlation = 0.01 [− 0.02; 0.04], *N* = 65 combinations, t-test *p* = 0.6) (SI Figure 8).

Because of the mixed evidence, to further investigate the possibility that the frequency of resistance follows the fluctuations in antibiotic use, we computed two additional partially independent indicators (SI Figure 9) [24]. These also suggested that antibiotic resistance follows temporal fluctuations in the use of antibiotics in the hospital sector, but not in the community.

We last looked at whether *sustained* trends in antibiotic consumption (computed as the Spearman correlation between calendar year and consumption level) predict the temporal trajectory of antibiotic resistance. In other words, we tested whether increasing/decreasing resistance trajectories are associated with increasing/decreasing consumption of the corresponding antibiotic in the corresponding country. Hospital antibiotic consumption rate was increasing on average for all resistance trends (Figure 6 B), but the increase in consumption was significantly higher for the increasing (s and ns) than decreasing (s and ns) categories (Wilcox test, *p* = 0.002). Again, no significant trend was found when considering community instead of hospital consumption (Wilcox test, *p* = 0.14).

All in all these analyses provide some evidence that the frequency of antibiotic resistance follows changes in the use of the antibiotic – either year-on-year fluctuations and/or sustained trends – in the hospital, but not community sector.

### 2.5 Case-study: few rising trajectories in previously high-consumption countries

Finally, we combined the insights above to further understand specific resistance trends. Some of the countries with the smallest proportion of increasing trajectories (Figure 4 and SI Figure 2), such as Greece, Italy, and France, were also the countries with highest levels of antibiotic use in the community in the early 2000s [22]. Confirming this observation, there was overall a significant negative correlation between antibiotic use in the community sector in 2000 and the proportion of increasing resistance trajectories (SI Figure 10C). Such trend was not observed for hospital consumption.

We hypothesised two non-mutually exclusive explanations for this intriguing finding. One possibility is that countries with high community antibiotic use in the early 2000s have taken effective measures to halt the increase of antibiotic resistance. We looked at whether high community antibiotic use in 2000 correlated with a decrease in total antibiotic consumption during the study period. This was true in the hospital sector, but not in the community (SI Figure 10A,B). Alternatively, as high antibiotic consumption leads to a faster rise in resistance, trajectories in high consumption countries are more likely to have reached a plateau either before or during the study period, while trajectories in lower consumption countries are still rising towards the plateau. This hypothesis requires that increasing trajectories reflect, at least in part, non-equilibrium dynamics. In support of this hypothesis, we found that countries with high community antibiotic consumption in 2000 had a faster rate of increase in the rising phase of the stabilising trajectories (Spearman’s rho: 0.71, *p* = 0.0014).

## 3 Discussion

Our analysis of resistance trajectories in Europe suggests that antibiotic resistance frequencies are not consistently increasing and instead often appear stable over two decades. This observation has been previously discussed in the context of *S. pneumoniae* [17, 18, 20] and, to a more limited extent, in other species [19]. Our work provides systematic evidence for stability and – notably – stabilisation of resistance in eight important bacterial species. However, a non-negligible minority of trajectories were categorised as either significantly increasing (17%) or significantly decreasing (17%). These trajectories could reflect either non-equilibrium dynamics or a changing equilibrium frequency in response to changing antibiotic use. While we cannot determine for certain which is the case for individual trajectories, our analysis suggests a mix of these scenarios.

There was considerable variability in trends by bacterial species, country, and antibiotic class. We were able to explain some of this variability. First, resistances were in part associated with the rate of use of the corresponding antibiotic. In addition to previous work showing a spatial correlation between median consumption and median resistance [22, 23], we evidenced the effect of antibiotic consumption on both the rise and the stable level of resistance. We also investigated the *temporal* correlation between antibiotic use and resistance frequency. Resistance was weakly associated with hospital – but not community – antibiotic consumption, both in terms of year-on-year fluctuations and sustained trends. Second, the species *E. coli* and *K. pneumoniae* were associated with a particularly large number of increasing or stabilising trajectories. This might be linked to the emergence and spread of the CTX-M beta-lactamase enzyme in the late 1990s and early 2000s [25]. Third, countries with high antibiotic consumption in 2000 [22] – e.g. Italy, Greece and France – saw fewer increasing resistance trajectories than other countries. This was not straightforwardly explained by a decrease in antibiotic use, but might be explained by these trajectories having stabilised earlier in these countries than in others.

A large part of the variability in resistance within and across countries remained unexplained. One reason could be that differences in host population structure, and heterogeneity in antibiotic consumption *within* this structure, can lead to different resistance frequencies for populations with the same average consumption [19]. A second explanation is the spillover of resistance from neighbouring countries: even small amounts of migration from other countries are sufficient to influence local levels of resistance, which would tend to weaken the spatial correlation across countries [18, 26]. Thirdly, the coupling of multiple resistance genes co-occurring on bacterial strains (‘linkage disequilibrium’) [27] means that the frequency of resistance against a particular antibiotic could be affected by the consumption of other antibiotics. Finally, a number of studies have reported statistical associations between the frequency of resistance and other factors – e.g. temperature, pollution, governance- and infrastructure-related factors [15, 28, 29]. While this strongly suggests selection pressure is not solely dependent on antibiotic consumption, it is challenging to determine which factors are truly causal. Determining the causal role of factors beyond average antibiotic consumption requires detailed epidemiological and genomic data, including host metadata (host characteristics, geographic location), contact networks, and sequence data conveying the genetic coupling between different resistances and other loci determining important bacterial phenotypes.

One important limitation of our study is data quality and consistency across countries. The tested isolates do not necessarily reflect a random sample of bacteria circulating in the human population, or even of bacteria infecting hospital patients. For example, intensified testing in the context of hospital outbreaks might cause the over-representation of particular clones in a specific country and year. Resistance frequencies are not comparable across years if the threshold for resistance is changed, as was the case in France for *P. aeruginosa* [30]. The antibiotic consumption data is subject to similar data quality considerations. To limit the inclusion of biased or poor-quality data in our analyses, we did not consider the trajectories for which none of our models fitted the data well (‘poor fit’ category), and we removed temporal outliers from the consumption data. Despite these concerns, most trajectories were quite smooth temporally (Figure 3). Furthermore, while potential biases will affect the overall estimate of resistance frequency, as long as bias remains constant in time, they will not affect the detection and categorisation of temporal trends. Although the data from the ECDC may represent one of the largest and best standardised datasets on antibiotic resistance and consumption, it is limited to European countries. It is important to determine whether the pattern of stability and stabilising also holds elsewhere, particularly in low and middle income countries (LMICs). While data sources covering a wider range of countries exist (e.g. ResistanceMap^1^ and ATLAS^2^), their temporal span is currently much smaller for LMICs than the two decades covered by the European network. Thus, while quantifying overall resistance trends in LMICs is possible [15], assessing evidence for stabilisation may prove more challenging.

Finally, the stability of resistance trends may not be robust to evolutionary innovation. With sustained selection pressure, bacteria may evolve new and less costly resistance mechanisms and compensatory mutations that alleviate the cost of resistance. This would result in the progressively increase the equilibrium level of resistance. While compensatory evolution is readily observed under laboratory conditions, its contribution to real-word resistance dynamics is unclear.

In conclusion, using antibiotic resistance data from 3,375,774 bacterial isolates from infections, encompassing 8 bacterial species, 30 European countries, and 20 years, we reveal that resistance trajectories are for the most part stable or stabilising. This overall behaviour of antibiotic resistance trajectories suggests that ever increasing antibiotic resistance is not inevitable and that resistance management can be effective in limiting resistance spread. Observed resistance dynamics were, to some extent, explainable by levels of antibiotic consumption. However, this explanatory power was relatively low, highlighting important gaps in our understanding of resistance dynamics. All in all, we open perspectives for further work elucidating how the complex ecology of bacteria determine the long-term fate of antibiotic resistance.

## Data Availability

The data is available upon request from the European Centre for Disease Prevention and Control (ECDC). All code is available on the following GitHub repository: https://github.com/mjemons/temporal-trends-AMR-manuscript

## Acknowledgements

FB is funded by ERC StG 949208 EvoComBac. SL is funded by the Swiss National Science Foundation (PR00P3 201618).

Data from The European Surveillance System—TESSy, provided by Finland, Sweden, Belgium, Germany, Greece, Ireland, Italy, Luxembourg, Netherlands, Norway, Portugal, United Kingdom, Austria, Bulgaria, Czech Republic, Denmark, Estonia, Spain, Malta, Slovenia, France, Croatia, Hungary, Poland, Slovakia, Romania Cyprus, Latvia, Lithuania and released by ECDC. The views and opinions of the authors expressed herein do not necessarily state or reflect those of ECDC. The accuracy of the authors’ statistical analysis and the findings they report are not the responsibility of ECDC. ECDC is not responsible for conclusions or opinions drawn from the data provided. ECDC is not responsible for the correctness of the data and for data management, data merging and data collation after provision of the data. ECDC shall not be held liable for improper or incorrect use of the data.

## 4 Methods

### 4.1 Data

#### 4.1.1 Resistance Raw Data

The data on the antimicrobial resistance rates are released by the European Surveillance System -TESSy, provided by Finland, Sweden, Belgium, Germany, Greece, Ireland, Italy, Luxembourg, Netherlands, Norway, Portugal, United Kingdom, Austria, Bulgaria, Czech Republic, Denmark, Estonia, Spain, Malta, Slovenia, France, Croatia, Hungary, Poland, Slovakia, Romania Cyprus, Latvia, Lithuania and released by the European Centre for Disease Prevention and Control (ECDC). The data covers the years 1998-2019.

The raw data, available upon request from the ECDC, is in case-based format, and consists of, for each isolate: the date of the isolation, the pathogen, the patient type (inpatient or outpatient), and the antimicrobial resistance level (sensitive, intermediate, resistant according to EUCAST clinical breakpoints) to various antibiotics. Outpatients consist of patients presenting at the hospital for dialysis, other day hospital care, or emergency. A more detailed description of the raw resistance data is made available by the ECDC [31].

#### 4.1.2 Antibiotic Consumption Raw Data

Data on antibiotic consumption is publicly available from the ECDC, and consists of defined daily dose (DDD) per 1000 inhabitants by antibiotic class, country, year, sector (community, hospital care, total care) and uptake route (oral, parenteral, rectal, inhalation, implant, all). The dataset includes the same 30 countries as the resistance dataset, spans the years 1997-2016, and consists of the following classes of antibiotics: J01A (tetracyclines), J01B (amphenicols), J01C (penicillins), J01D (other betalactams), J01E (sulfonamides), J01F (macrolides), J01G (aminoglycosides), J01M (quinolones), J01R (combinations of antibacterials), J01X (others), P01A (agents against amoebiasis and other protozoal diseases), A07A (intestinal antiinfectives). A detailed description of the consumption data is made available by the ECDC [32].

### 4.2 Preprocessing

#### 4.2.1 Processing of antibiotic resistance data

We computed the frequency of resistant for each bug, drug, country, year, and patient type combination. Intermediate isolates were considered resistant. We compared the temporal trends in resistance in inpatients and outpatients for drug-pathogen-country combinations that had data on both types of patients. The temporal trends in outpatients and in patients were very correlated, with similar or higher levels of resistance in inpatients. Therefore we only considered resistance measured in inpatients for further analyses.

#### 4.2.2 List of the bug-drug combinations investigated

*S*.*aureus*: CIP CLO DAP DIC FLC FOX LNZ LVX MET NOR OFX OXA RIF TEC

*S. pneumoniae:* AZM CIP CLR CRO CTX ERY LVX MFX NOR OFX OXA PEN

E. faecalis: AMP AMX GEH LNZ TEC VAN

*E. faecium:* AMP AMX GEH LNZ TEC VAN

*E. coli* : AMC AMK AMP AMX CAZ CIP COL CRO CTX DOR ETP FEP GEN IPM LVX MEM MFX NAL NET NOR OFX PIP POL TGC TOB TZP

*K. pneumoniae:* AMC AMK CAZ CIP COL CRO CTX DOR ETP FEP GEN IPM LVX MEM MFX NAL NET NOR OFX PIP POL TGC TOB TZP

*P. aeruginosa:* AMK CAZ CIP COL DOR ETP FEP GEN IPM LVX MEM NET PIP POL TOB TZP

*Acinetobacter spp*.: AMC AMK AMP AMX CAZ CIP COL CTX DOR FOX GEH GEN IPM LVX MEM NAL NET POL TOB TZP VAN

#### 4.2.3 Processing of antibiotic use data

For many countries and years, the total consumption of antibiotics by all routes of administration was reported. We primarily used this information when it was available. Moreover, some countries and years recorded the consumption decomposed by route of administration of antibiotics. This was most commonly oral and parenteral, and, less frequently, inhalation, rectal, implant, “other” routes. When total consumption was not available, we summed the consumption recorded over all routes to obtain an estimate of total consumption.

We considered consumption both in the hospital sector and in the community.

To remove all outliers for a given antibiotic class, country, sector, route of administration for each combination we removed datapoints for years not within *±*3 *×IQR* (IQR = Inter quartile range) of the temporal median.

### 4.3 Inferring temporal trends in antibiotic resistance

For combinations of bug, drug, country, we fitted logistic models to the temporal trends in the frequency of resistance. We only included the combinations with a time series of at least 5 years, where all years had at least 30 isolates tested for drug resistance, and in total at least 10 resistant bacterial isolates. Fitting was done with non-linear least square regression, with the function `nlsLM` from the `minpack.lm` R package, except for the flat line which was fit using `lm` from the base `stats` package [33, 34]. The error is assumed to be normally distributed and proportional to the inverse of the sample size for each year.

We fitted three models to the temporal trends data:

1. A standard logistic model with two parameters, an offset and a slope. The standard logistic model results in an increasing sigmoid from 0 to 1, or a decreasing sigmoid from 1 to 0 if the slope is negative. This corresponds to the standard population genetics model describing the action of selection on resistance frequency. The resistance frequency *f* for year *t* under this model is given by:

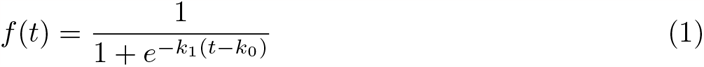

where *k*_1_ is the slope parameter and *k*_0_ determines the offset.
2. A ‘flat’ model with slope 0 and an intercept. In this model, the resistance frequency *f* for year *t* is given by:

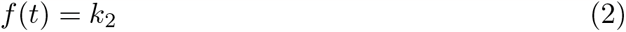
3. A plateauing logistic model with a maximum frequency below 1, where the sigmoid function is scaled by parameter *k*_2_. In this model, the resistance frequency *f* for year *t* is given by:

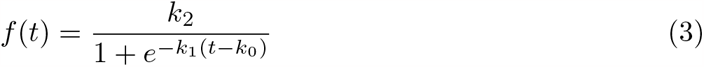

We compared the models using the corrected Akaike Information Criterion (AICc), with the best model corresponding to the smallest AICc [35]. The AICc is given by:

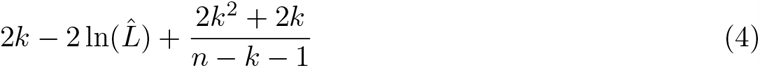

where 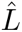 is the maximum likelihood estimate, *k* the number of parameters and *n* the sample size.

We quantified the goodness of fit with the mean absolute error of the best fit model. For the two logistic models, we determined the statistical significance of the slope based on a threshold of *p <* 0.05.

This resulted in six categories of temporal trends:

i. significantly increasing, when the best model was the standard logistic and the slope was significantly positive.
ii. non-significantly increasing, when the best model was the standard logistic and the slope was positive but not significantly different from 0
iii. non-significantly decreasing, when the best model was logistic and the slope was negative but not significantly different from 0.
iv. significantly decreasing, when the best model was logistic and the slope was significantly negative.
v. stable, when the best model was a flat fit.
vi. stabilising, when the best model was the logistic with plateau different from 1.

Additionally, some trajectories were classified as ‘poor fit’ when the mean absolute error exceeded 0.05 and were not analysed further. The ‘stabilising’ category can be subdivided further in ‘stabilising significantly increasing’ and ‘stabilising non-significantly increasing’ depending on the significance of the slope leading to the plateau.

### 4.4 Comparison of rate of resistance change across models

The slope parameters in the standard and scaled logistic models (*k*_1_ in Equations 1 and 3) are not directly comparable. This is because the parameter reflects how quickly resistance will reach its maximum level. As the maximum is different in the two models, the same slope parameter in the two models translates to a different rate of change in the actual resistance frequencies. For the analyses in which we sought to compare the speed of change across the two models i.e. the comparison of increasing and stabilising trajectories and the correlation with antibiotic usage – we computed a comparable measure of the change in resistance frequencies. Specifically, we looked at the maximal rate of change observed in the temporal window for which we had data.

### 4.5 Correlation between antimicrobial consumption and resistance

We correlated antimicrobial consumption and resistance across countries and time. In all cases, we correlated the level of resistance to a particular antibiotic to the total consumption of all antibiotics of this class.

#### 4.5.1 Correlation between antimicrobial consumption and resistance across countries

For each bug-drug combination, we correlated across countries the median antibiotic consumption across years, to three properties of the resistance temporal trends. These three properties were the median resistance across years, the resistance plateau (for ‘stable’ and ‘stabilising’ categories), the slope of the logistic (for ‘logistic significantly increasing’ and ‘stabilising significantly increasing’ categories). As described above, the slope was computed as the maximal rate of change reached in the temporal window for which we had data This ensures the slope is comparable across the logistic model with and without intermediate plateau, unlike the slope parameter of the logistic. We quantified the correlation with Spearman’s rank correlation coefficient across countries, for bug-drug combinations represented by at least five countries. The correlation was calculated using `SpearmanRho` from `DescTools version 0.99.49` [36].

This analysis resulted in a number of correlation coefficients. These coefficients are calculated for combinations of antibiotics prescribed in the community vs. the hospital sector, (we remind that we consider only resistance in inpatients), and of the three properties of temporal trends which are correlated (total of 6 combinations).

#### 4.5.2 Correlation between antimicrobial consumption and resistance across time

For each country-bug-drug combination, we correlated across years the level of resistance and level of antibiotic consumption. This results in a Spearman’s rank correlation coefficient for the country-bug-drug combinations and each of the four combinations of community vs. hospital sector. This was again calculated using `SpearmanRho` from `DescTools version 0.99.49` [36]. We kept only country-bug-drug combinations with at least 5 years of data available. We averaged these correlations over countries for each bug-drug combination, retaining only bug-drug combinations for which 5 countries of more were represented. It is expected that the temporal correlation between the frequency of resistance and the corresponding antibiotic use is positive if the bacterial population adapts to the changing use of specific antibiotic.

We additionally computed a cross-temporal correlation, whereby the consumption data were shifted by -1 and +1 year compared to the resistance data. To give additional indications that yearly variation in antibiotic resistance follows variation in consumption, we used these cross-temporal correlations. First, if the bacterial population adapts closely to the contemporaneous antibiotic use, it is expected that the “temporal adaptation” pattern formed by the cross-correlation for time-shifts − 1, 0 and +1 is maximal at 0 [24]. We counted the fraction of bug-drug combinations for which temporal adaptation was maximal at 0 and calculated the probability of this fraction under a binomial model with true probability 0.25. Second, we compared the temporal correlation with the spatial correlation across bug-drug combinations. Indeed, if the bacterial population evolves in response to changing antibiotic use, it is expected that levels of resistance match both the contemporaneous and local antibiotic use [24]. We therefore tested whether bug-drug combinations with stronger spatial correlations also exhibited stronger temporal correlations, by correlating the two correlations across bug-drug combinations (Figure 9).

### 4.6 Code Availability

All code is available on the following GitHub repository:

https://github.com/mjemons/temporal-trends-AMR-manuscript

All analyses were performed with `R version 4.2.3` [34]. Packages are managed with `renv` [37].

## Footnotes

1 https://resistancemap.onehealthtrust.org/AntibioticResistance.php

2 https://atlas-surveillance.com

## References

[1] Christopher JL Murray, Kevin Shunji Ikuta, Fablina Sharara, Lucien Swetschinski, Gisela Robles Aguilar, Authia Gray, Chieh Han, Catherine Bisignano, Puja Rao, Eve Wool, Sarah C. Johnson, Annie J. Browne, Michael Give Chipeta, Frederick Fell, Sean Hackett, Georgina Haines-Woodhouse, Bahar H. Kashef Hamadani, Emmanuelle A. P. Kumaran, Barney McManigal, Ramesh Agarwal, Samuel Akech, Samuel Albertson, John Amuasi, Jason Andrews, Aleskandr Aravkin, Elizabeth Ashley, Freddie Bailey, Stephen Baker, Buddha Basnyat, Adrie Bekker, Rose Bender, Adhisivam Bethou, Julia Bielicki, Suppawat Boonkasidecha, James Bukosia, Cristina Carvalheiro, Carlos Castañeda-Orjuela, Vilada Chansamouth, Suman Chaurasia, Sara Chiurchiù, Fazle Chowdhury, Aislinn J. Cook, Ben Cooper, Tim R. Cressey, Elia Criollo-Mora, Matthew Cunningham, Saffiatou Darboe, Nicholas P. J. Day, Maia De Luca, Klara Dokova, Angela Dramowski, Susanna J. Dunachie, Tim Eckmanns, Daniel Eibach, Amir Emami, Nicholas Feasey, Natasha Fisher-Pearson, Karen Forrest, Denise Garrett, Petra Gastmeier, Ababi Zergaw Giref, Rachel Claire Greer, Vikas Gupta, Sebastian Haller, Andrea Haselbeck, Simon I. Hay, Marianne Holm, Susan Hopkins, Kenneth C. Iregbu, Jan Jacobs, Daniel Jarovsky, Fatemeh Javanmardi, Meera Khorana, Niranjan Kissoon, Elsa Kobeissi, Tomislav Kostyanev, Fiorella Krapp, Ralf Krumkamp, Ajay Kumar, Hmwe Hmwe Kyu, Cherry Lim, Direk Limmathurotsakul, Michael James Loftus, Miles Lunn, Jianing Ma, Neema Mturi, Tatiana Munera-Huertas, Patrick Musicha, Marisa Marcia Mussi-Pinhata, Tomoka Nakamura, Ruchi Nanavati, Sushma Nangia, Paul Newton, Chanpheaktra Ngoun, Amanda Novotney, Davis Nwakanma, Christina W. Obiero, Antonio Olivas-Martinez, Piero Olliaro, Ednah Ooko, Edgar Ortiz-Brizuela, Anton Yariv Peleg, Carlo Perrone, Nishad Plakkal, Alfredo Ponce-de Leon, Mathieu Raad, Tanusha Ramdin, Amy Riddell, Tamalee Roberts, Julie Victoria Robotham, Anna Roca, Kristina E. Rudd, Neal Russell, Jesse Schnall, John Anthony Gerard Scott, Madhusudhan Shivamallappa, Jose Sifuentes-Osornio, Nicolas Steenkeste, Andrew James Stewardson, Temenuga Stoeva, Nidanuch Tasak, Areerat Thaiprakong, Guy Thwaites, Claudia Turner, Paul Turner, H. Rogier van Doorn, Sithembiso Velaphi, Avina Vongpradith, Huong Vu, Timothy Walsh, Seymour Waner, Tri Wangrangsimakul, Teresa Wozniak, Peng Zheng, Benn Sartorius, Alan D. Lopez, Andy Stergachis, Catrin Moore, Christiane Dolecek, and Mohsen Naghavi. Global burden of bacterial antimicrobial resistance in 2019: a systematic analysis. The Lancet, 399(10325):629–655, February 2022. ISSN 0140-6736, 1474-547X. doi: 10.1016/S0140-6736(21)02724-0. URL https://www.thelancet.com/journals/lancet/article/PIIS0140-6736(21)02724-0/fulltext. Publisher: Elsevier.

[2] Christopher Witzany, Sebastian Bonhoeffer, and Jens Rolff. Is antimicrobial resistance evolution accelerating? PLOS Pathogens, 16(10):e1008905, October 2020. ISSN 1553-7374. doi: 10.1371/journal.ppat.1008905. URL https://journals.plos.org/plospathogens/article?id=10.1371/journal.ppat.1008905. Publisher: Public Library of Science.

[3] Althea W McCormick, Cynthia G Whitney, Monica M Farley, Ruth Lynfield, Lee H Harrison, Nancy M Bennett, William Schaffner, Arthur Reingold, James Hadler, Paul Cieslak, et al. Geographic diversity and temporal trends of antimicrobial resistance in streptococcus pneumoniae in the united states. Nature medicine, 9(4):424–430, 2003.

[4] L. Clifford McDonald. Trends in Antimicrobial Resistance in Health Care–Associated Pathogens and Effect on Treatment. Clinical Infectious Diseases, 42(Supplement 2):S65–S71, January 2006. ISSN 1058-4838. doi: 10.1086/499404. URL https://doi.org/10.1086/499404.

[5] Lotta Siira, Merja Rantala, Jari Jalava, Antti J. Hakanen, Pentti Huovinen, Tarja Kaijalainen, Outi Lyytikäinen, and Anni Virolainen. Temporal trends of antimicrobial resistance and clonality of invasive Streptococcus pneumoniae isolates in Finland, 2002 to 2006. Antimicrobial Agents and Chemotherapy, 53(5):2066–2073, May 2009. ISSN 1098-6596. doi: 10.1128/AAC.01464-08.

[6] A. Fenoll, J. J. Granizo, L. Aguilar, M. J. Giménez, L. Aragoneses-Fenoll, G. Hanquet, J. Casal, and D. Tarragó. Temporal trends of invasive Streptococcus pneumoniae serotypes and antimicrobial resistance patterns in Spain from 1979 to 2007. Journal of Clinical Microbiology, 47(4):1012–1020, April 2009. ISSN 1098-660X. doi: 10.1128/JCM.01454-08.

[7] Majdi N Al-Hasan, Brian D Lahr, Jeanette E Eckel-Passow, and Larry M Baddour. Antimicrobial resistance trends of escherichia coli bloodstream isolates: a population-based study, 1998–2007. Journal of antimicrobial chemotherapy, 64(1):169–174, 2009.

[8] Robert D Kirkcaldy, Sarah Kidd, Hillard S Weinstock, John R Papp, and Gail A Bolan. Trends in antimicrobial resistance in neisseria gonorrhoeae in the usa: the gonococcal isolate surveillance project (gisp), january 2006–june 2012. Sexually transmitted infections, 89 (Suppl 4):iv5–iv10, 2013.

[9] F.-P. Hu, Y. Guo, D.-M. Zhu, F. Wang, X.-F. Jiang, Y.-C. Xu, X.-J. Zhang, C.-X. Zhang, P. Ji, Y. Xie, M. Kang, C.-Q. Wang, A.-M. Wang, Y.-H. Xu, J.-L. Shen, Z.-Y. Sun, Z.- J. Chen, Y.-X. Ni, J.-Y. Sun, Y.-Z. Chu, S.-F. Tian, Z.-D. Hu, J. Li, Y.-S. Yu, J. Lin, B. Shan, Y. Du, Y. Han, S. Guo, L.-H. Wei, L. Wu, H. Zhang, J. Kong, Y.-J. Hu, X.-M. Ai, C. Zhuo, D.-H. Su, Q. Yang, B. Jia, and W. Huang. Resistance trends among clinical isolates in China reported from CHINET surveillance of bacterial resistance, 2005-2014. Clinical Microbiology and Infection: The Official Publication of the European Society of Clinical Microbiology and Infectious Diseases, 22 Suppl 1:S9–14, March 2016. ISSN 1469-0691. doi: 10.1016/j.cmi.2016.01.001.

[10] Dee Shortridge, Ana C. Gales, Jennifer M. Streit, Michael D. Huband, Athanasios Tsakris, and Ronald N. Jones. Geographic and Temporal Patterns of Antimicrobial Resistance in Pseudomonas aeruginosa Over 20 Years From the SENTRY Antimicrobial Surveillance Program, 1997-2016. Open Forum Infectious Diseases, 6(Suppl 1):S63–S68, March 2019. ISSN 2328-8957. doi: 10.1093/ofid/ofy343.

[11] Jean Carlet, Vincent Jarlier, Jacques Acar, Olivier Debaere, Patrick Dehaumont, Bruno Grandbastien, Pierre Le Coz, Gerard Lina, Yves Pean, Claude Rambaud, France Roblot, Jérôme Salomon, Benoit Schlemmer, Pierre Tattevin, and Benoit Vallet. Trends in Antibiotic Consumption and Resistance in France Over 20 Years: Large and Continuous Efforts but Contrasting Results. Open Forum Infectious Diseases, 7(11):ofaa452, November 2020. ISSN 2328-8957. doi: 10.1093/ofid/ofaa452.

[12] Rik Oldenkamp, Constance Schultsz, Emiliano Mancini, and Antonio Cappuccio. Filling the gaps in the global prevalence map of clinical antimicrobial resistance. Proceedings of the National Academy of Sciences, 118(1):e2013515118, 2021.

[13] Shirin Aliabadi, Elita Jauneikaite, Berit Müller-Pebody, Russell Hope, Karina-Doris Vihta, Carolyne Horner, and Céire E. Costelloe. Exploring temporal trends and risk factors for resistance in Escherichia coli-causing bacteraemia in England between 2013 and 2018: an ecological study. The Journal of Antimicrobial Chemotherapy, 77(3):782–792, February 2022. ISSN 1460-2091. doi: 10.1093/jac/dkab440.

[14] European Centre for Disease Prevention, Control, and World Health Organization. Antimicrobial resistance surveillance in europe 2023 - 2021 data. Stockholm, 2023.

[15] Eve Rahbe, Laurence Watier, Didier Guillemot, Philippe Glaser, and Lulla Opatowski. Determinants of worldwide antibiotic resistance dynamics across drug-bacterium pairs: a multivariable spatial-temporal analysis using atlas. The Lancet Planetary Health, 7(7): e547–e557, 2023.

[16] Marc Lipsitch. The rise and fall of antimicrobial resistance. Trends in Microbiology, 9(9): 438–444, September 2001. ISSN 0966-842X. doi: 10.1016/S0966-842X(01)02130-8. URL https://www.sciencedirect.com/science/article/pii/S0966842X01021308.

[17] Caroline Colijn, Ted Cohen, Christophe Fraser, William Hanage, Edward Goldstein, Noga Givon-Lavi, Ron Dagan, and Marc Lipsitch. What is the mechanism for persistent coexistence of drug-susceptible and drug-resistant strains of Streptococcus pneumoniae? Journal of The Royal Society Interface, 7(47):905–919, June 2010. doi: 10.1098/rsif.2009.0400. URL https://royalsocietypublishing.org/doi/full/10.1098/rsif.2009.0400. Publisher: Royal Society.

[18] François Blanquart, Sonja Lehtinen, Marc Lipsitch, and Christophe Fraser. The evolution of antibiotic resistance in a structured host population. Journal of The Royal Society Interface, 15(143):20180040, June 2018. doi: 10.1098/rsif.2018.0040. URL https://royalsocietypublishing.org/doi/full/10.1098/rsif.2018.0040. Publisher: Royal Society.

[19] Madison S. Krieger, Carson E. Denison, Thayer L. Anderson, Martin A. Nowak, and Alison L. Hill. Population structure across scales facilitates coexistence and spatial heterogeneity of antibiotic-resistant infections. PLOS Computational Biology, 16(7):e1008010, July 2020. ISSN 1553-7358. doi: 10.1371/journal.pcbi.1008010. URL https://journals.plos.org/ploscompbiol/article?id=10.1371/journal.pcbi.1008010. Publisher: Public Library of Science.

[20] Sonja Lehtinen, François Blanquart, Nicholas J. Croucher, Paul Turner, Marc Lipsitch, and Christophe Fraser. Evolution of antibiotic resistance is linked to any genetic mechanism affecting bacterial duration of carriage. Proceedings of the National Academy of Sciences, 114 (5):1075–1080, January 2017. ISSN 0027-8424, 1091-6490. doi: 10.1073/pnas.1617849114. URL https://www.pnas.org/content/114/5/1075. Publisher: National Academy of Sciences Section: Biological Sciences.

[21] Nicholas G. Davies, Stefan Flasche, Mark Jit, and Katherine E. Atkins. Within-host dynamics shape antibiotic resistance in commensal bacteria. Nature Ecology & Evolution, 3(3):440–449, March 2019. ISSN 2397-334X. doi: 10.1038/s41559-018-0786-x. URL https://www.nature.com/articles/s41559-018-0786-x. Number: 3 Publisher: Nature Publishing Group.

[22] Herman Goossens, Matus Ferech, Robert Vander Stichele, Monique Elseviers, and ESAC Project Group. Outpatient antibiotic use in Europe and association with resistance: a cross-national database study. Lancet (London, England), 365(9459):579–587, February 2005. ISSN 1474-547X. doi: 10.1016/S0140-6736(05)17907-0.

[23] Scott W Olesen, Michael L Barnett, Derek R MacFadden, John S Brownstein, Sonia Hernández-Díaz, Marc Lipsitch, and Yonatan H Grad. The distribution of antibiotic use and its association with antibiotic resistance. eLife, 7:e39435, December 2018. ISSN 2050-084X. doi: 10.7554/eLife.39435. URL https://doi.org/10.7554/eLife.39435. Publisher: eLife Sciences Publications, Ltd.

[24] François Blanquart and Sylvain Gandon. Time-shift experiments and patterns of adaptation across time and space. Ecology letters, 16(1):31–38, 2013.

[25] Paul-Louis Woerther, Charles Burdet, Elisabeth Chachaty, and Antoine Andremont. Trends in human fecal carriage of extended-spectrum β-lactamases in the community: toward the globalization of ctx-m. Clinical microbiology reviews, 26(4):744–758, 2013.

[26] Scott W Olesen, Marc Lipsitch, and Yonatan H Grad. The role of “spillover” in antibiotic resistance. Proceedings of the National Academy of Sciences, 117(46):29063–29068, 2020.

[27] Sonja Lehtinen, François Blanquart, Marc Lipsitch, Christophe Fraser, and with the Maela Pneumococcal Collaboration. On the evolutionary ecology of multidrug resistance in bacteria. PLoS pathogens, 15(5):e1007763, 2019.

[28] Peter Collignon, John J Beggs, Timothy R Walsh, Sumanth Gandra, and Ramanan Laxminarayan. Anthropological and socioeconomic factors contributing to global antimicrobial resistance: a univariate and multivariable analysis. The Lancet Planetary Health, 2(9): e398–e405, 2018.

[29] Zhenchao Zhou, Xinyi Shuai, Zejun Lin, Xi Yu, Xiaoliang Ba, Mark A Holmes, Yonghong Xiao, Baojing Gu, and Hong Chen. Association between particulate matter (pm) 2·5 air pollution and clinical antibiotic resistance: a global analysis. The Lancet Planetary Health, 7(8):e649–e659, 2023.

[30] C Slekovec, J Robert, Denis Trystram, JM Delarbre, A Merens, N van Der Mee-Marquet, C de Gialluly, Y Costa, J Caillon, D Hocquet, et al. Pseudomonas aeruginosa in french hospitals between 2001 and 2011: back to susceptibility. European journal of clinical microbiology & infectious diseases, 33:1713–1717, 2014.

[31] ECDC. TESSy-The European Surveillance System Antimicrobial resistance (AMR) reporting protocol 2021 European Antimicrobial Resistance Surveillance Network (EARS-Net) surveillance data for 2020. (February):38, 2021. URL https://www.ecdc.europa.eu/sites/default/files/documents/EARS-Net-reporting-protocol-2021_v2.pdf.

[32] ECDC. TESSy -The European Surveillance System Antimicrobial consumption (AMC) reporting protocol 2022 European Surveillance of Antimicrobial Consumption Network (ESAC-Net) surveillance data for 2022. (March), 2022. URL https://www.ecdc.europa.eu/sites/default/files/documents/ESACNet_protocol_2022.pdf.

[33] Timur V Elzhov, Katharine M Mullen, Andrej-Nikolai Spiess, and Ben Bolker Maintainer. minpack.lm: R Interface to the Levenberg-Marquardt Nonlinear Least-Squares Algorithm Found in MINPACK, Plus Support for Bounds. R package version 1.2-1, pages 1–14, 2016. URL https://cran.r-project.org/web/packages/minpack.lm/minpack.lm.pdf.

[34] R Core Team. R: A Language and Environment for Statistical Computing. R Foundation for Statistical Computing, Vienna, Austria, 2022. URL https://www.R-project.org/.

[35] Kenneth P Burnham and David R Anderson. Multimodel inference: understanding aic and bic in model selection. Sociological methods & research, 33(2):261–304, 2004.

[36] Andri Signorell, Ken Aho, Andreas Alfons, Nanina Anderegg, Tomas Aragon, Antti Arppe, Adrian Baddeley, Kamil Barton, Ben Bolker, Hans W Borchers, et al. Desctools: Tools for descriptive statistics. R package version 0.99, 28:17, 2019.

[37] Kevin Ushey. renv: Project Environments, 2023. URL https://CRAN.R-project.org/package=renv. R package version 0.17.3.

