## Supporting Information for "Temporal Trends in Antibiotic Resistance in Europe, 1998-2019"

September 27, 2023

#### 1 Categorisation of temporal trends

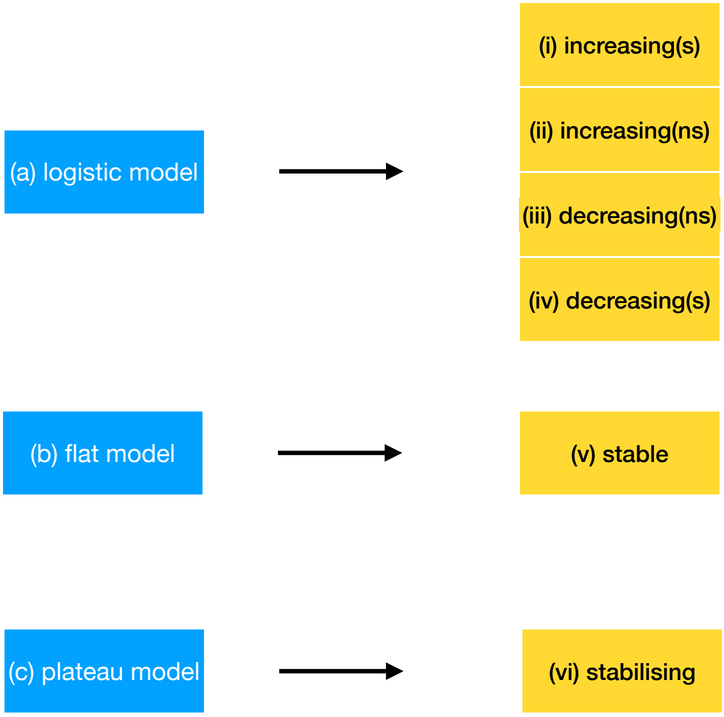


Figure 1: **Model fitting and categorisation of temporal trends**. This diagram shows schematically how the process of model fitting works. The three different models a-c are represented by the blue boxes. The categories that are created from the models a-c are represented by the yellow boxes i-vi.

#### 2 Predictors of temporal trends

To reduce the complexity of the analysis and make it easier to interpret, we first combined our categories into 'rising' (increasing (s) and (ns)), 'declining' (decreasing (s) and (ns)) and 'equilibrium' (stable and stabilising) trends. We then fitted a separate binomial regression for each combined category (i.e. 'rising' vs not as the outcome variable, 'declining' vs not declining, equilibrium vs not equilibrium) using `glm` from `R stats version 3.6.2`, with species, country and antibiotic class as predictor variables (SI Figure 2). For each regression, we used *Acinetobacter spp.* as the reference species, Austria as the reference country and J01A as the reference antibiotic class.

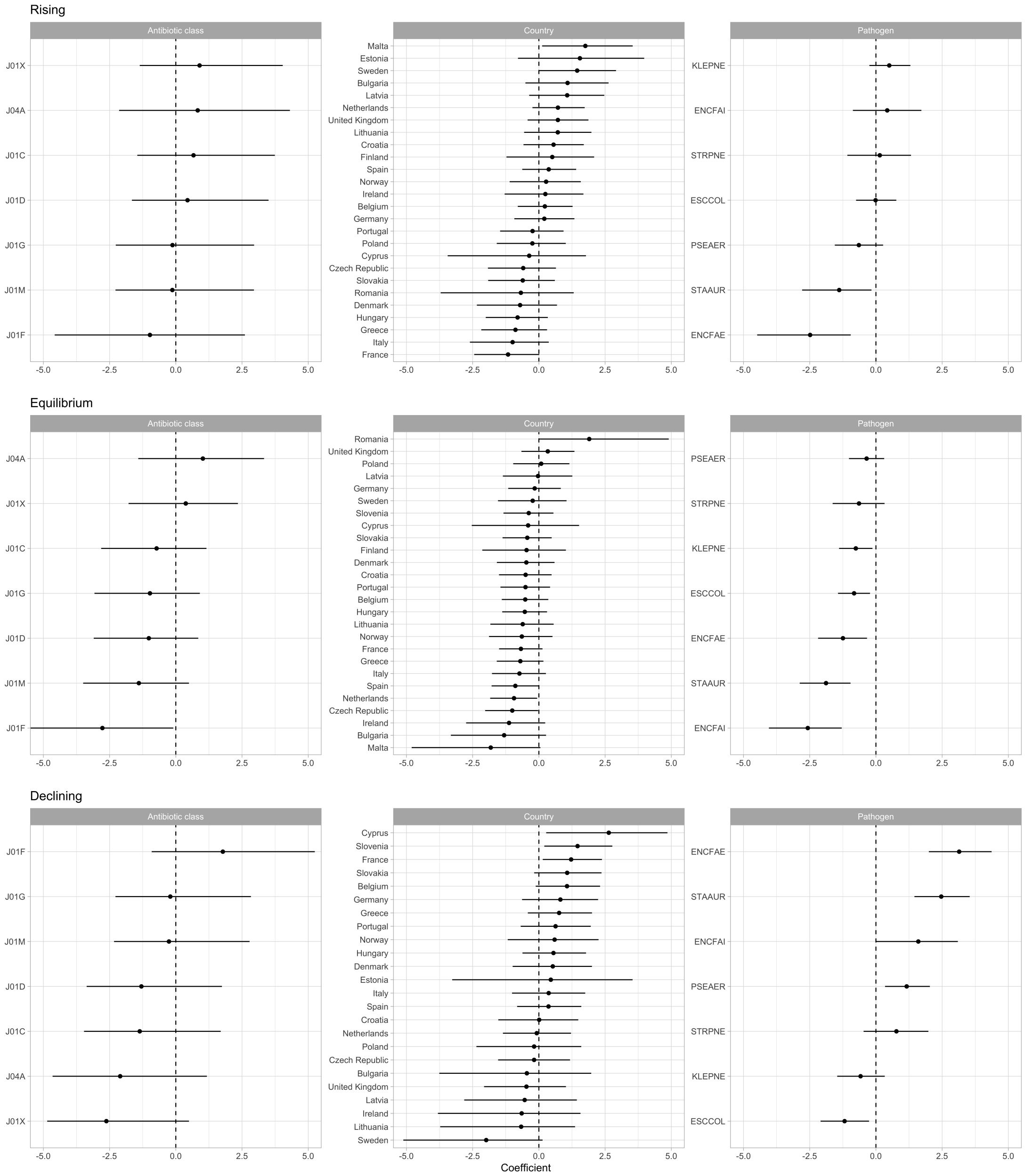


Figure 2: **Predictors of each type of temporal trend (i.e. rising, declining, and equilibrium).** The x-axis represents the coefficient of a binomial regression with species, country and antibiotic class as predictors of temporal trend (e.g. rising vs others). The reference species (for which the effect size is 0) is *Acinetobacter spp.*. The reference country is Austria. The reference antibiotic class is J01A. Error bars show 95% confidence intervals. In a small number of cases, countries had very little variability in temporal trends (e.g. no rising trajectories detected in Slovenia and Luxembourg); this results in an extreme value for the country's coefficient and very high uncertainty. We have therefore omitted these cases from the plots.

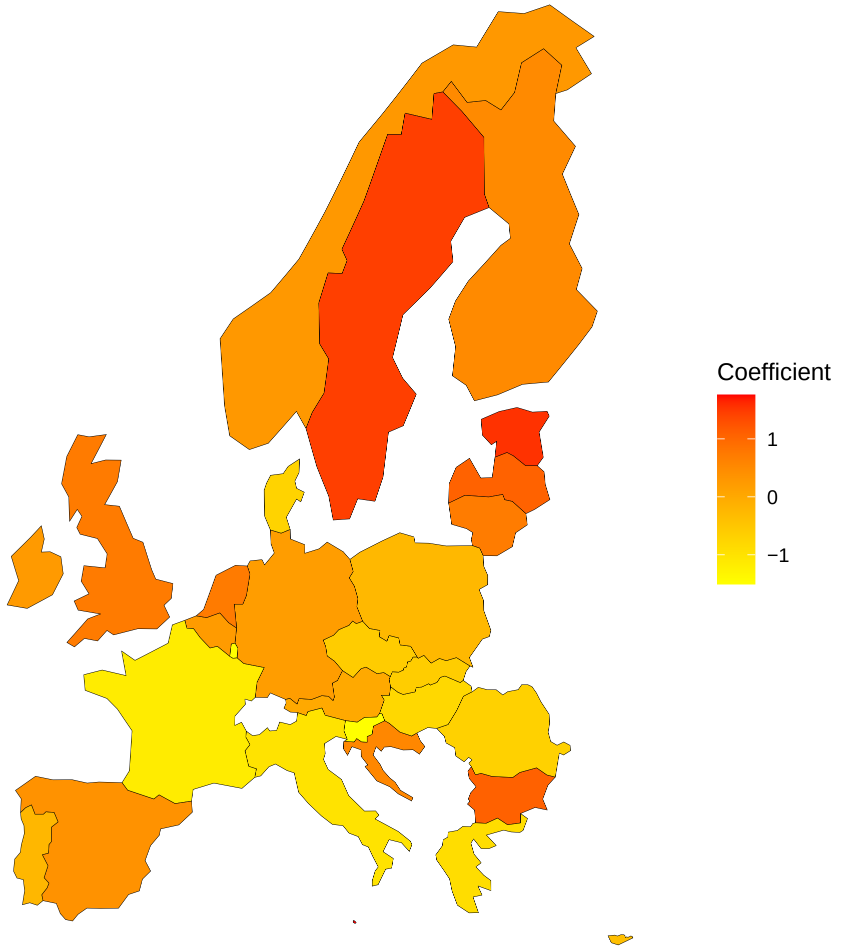


Figure 3: **Predictors of rising vs. not rising as depicted on a map of Europe.** A map of Europe coloured by the coefficient of the binomial regression. The reference country is Austria (coefficient equal to 0). Slovenia and Luxembourg do not present any increasing trend, and were associated with an extremely negative coefficient of -16 in the binomial regression; for visualisation purpose, we set the coefficient of these countries to -1.5

### 3 Comparison of rate of change of resistance frequencies

In the main text, we compared the rate at which resistance rises in the 'increasing (s)' and the non-equilibrium phase of the 'stabilising' categories. We found the mean rate to be smaller in the 'increasing (s)' trajectory and interpreted this as suggesting some of the increasing trajectories represent a changing equilibrium frequency rather than non-equilibrium dynamics. This interpretation depends on two assumptions. The first assumption is that the rising phase of the stabilising trajectories reflects genuine non-equilibrium dynamics, i.e. the trajectory from the initial emergence of the resistance to its equilibrium frequency. The second assumption is that a changing equilibrium leads to slower changes than non-equilibrium dynamics. This assumption arises because year-on-year variation in antibiotic usage is small compared to the actual level of antibiotic usage, which determines the speed of increase after initial emergence in the stabilising trajectories.

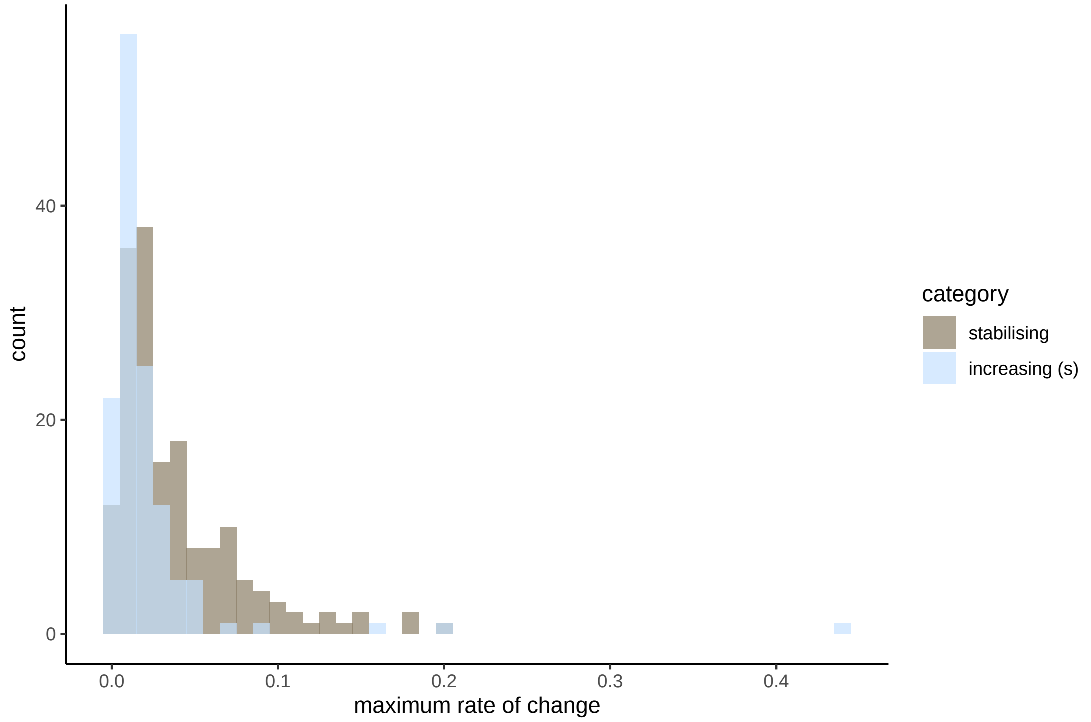


Figure 4: Comparison of the speed of change of resistance frequencies in the significantly increasing and stabilising categories. The speed of change is quantified as the maximum rate of change in the time window we have data for. The lower rate of change in the increasing category suggest that the increase may reflect a changing equilibrium rather than non-equilibrium dynamics.

### 4 Link between resistance and antibiotic consumption

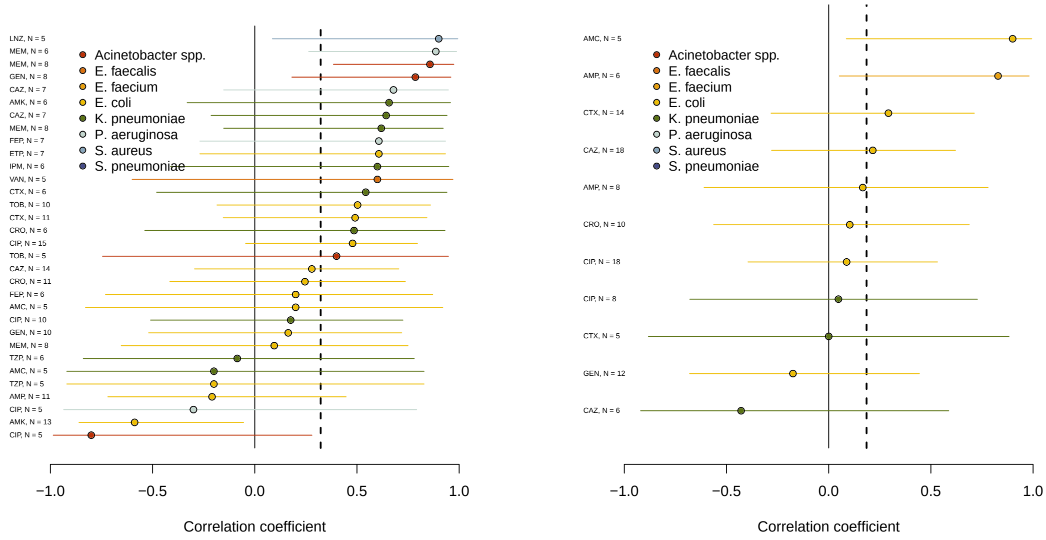


Figure 5: Spatial correlation coefficients between the **plateau** frequency of antibiotic resistance (left), or the **slope** (right), and the rate of use of the corresponding antibiotic in **hospitals**, for all bug-drug combinations. The number of countries included is indicated for each combination. The vertical dashed lines show the overall mean.

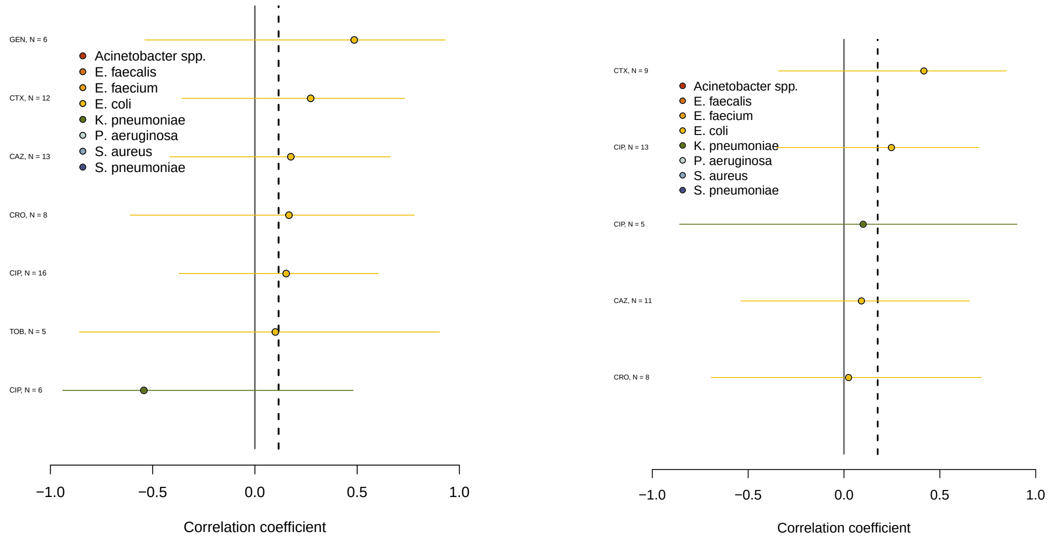


Figure 6: Spatial correlation coefficients between the rate of increase of antibiotic resistance, and the rate of use of the corresponding antibiotic in the community (left) or in the hospital (right), for all bug-drug combinations, when **restricting to stabilising temporal trajectories**. The number of countries included is indicated for each combination. The vertical dashed lines show the overall mean.

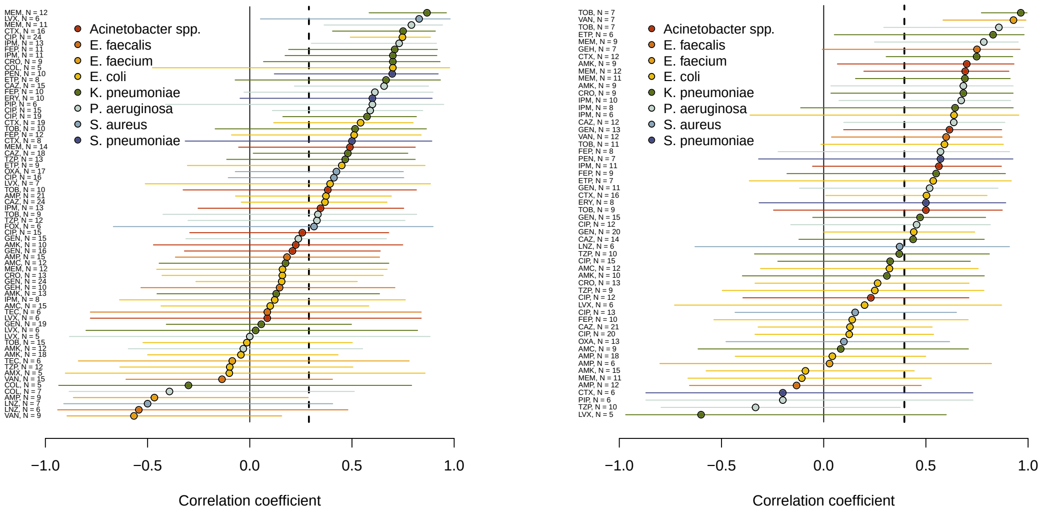


Figure 7: Spatial correlation coefficients between the **median** frequency of antibiotic resistance and the rate of use of the corresponding antibiotic in the **community** (left), or in the **hospitals** (right), for all bug-drug combinations. The number of countries included is indicated for each combination. The vertical dashed lines show the overall mean.

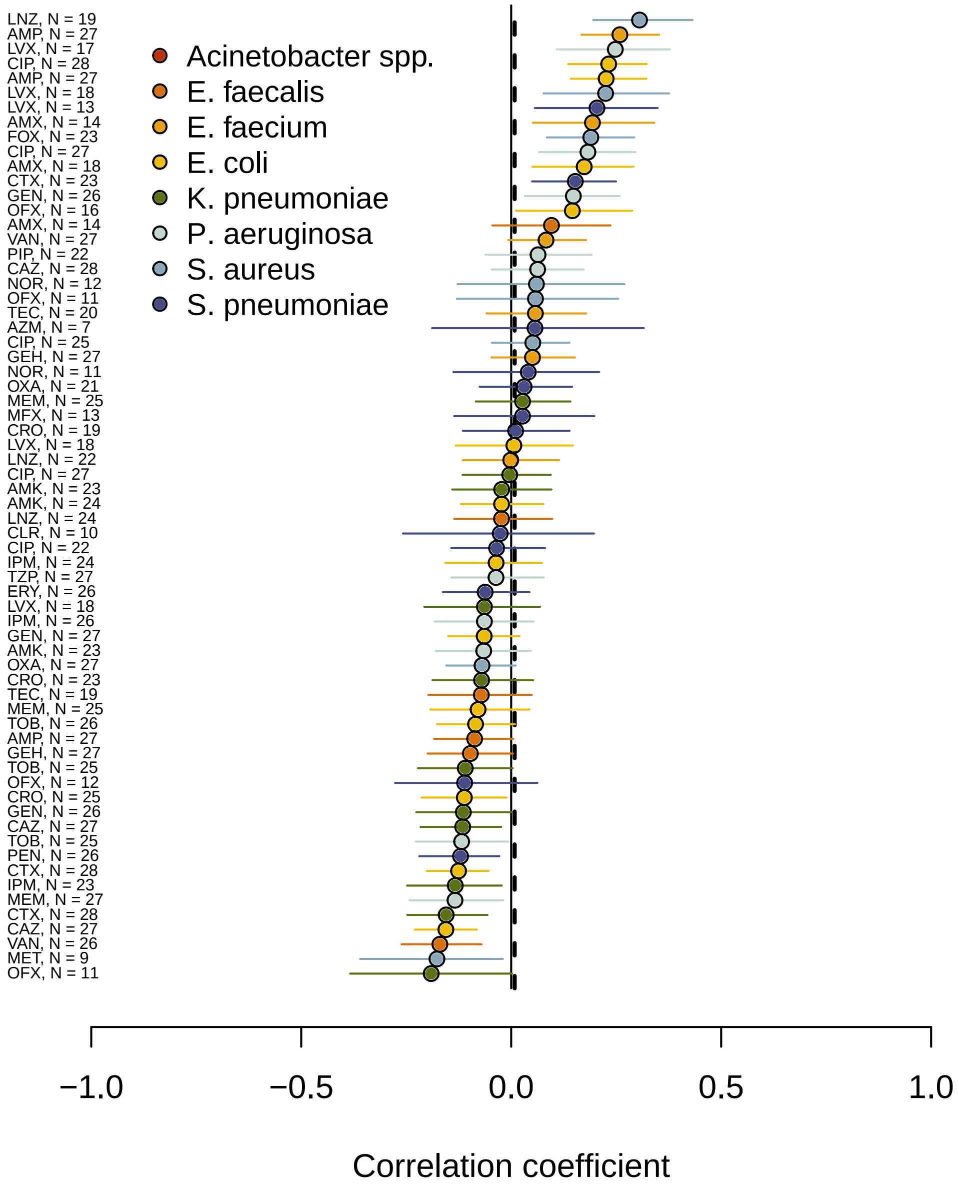


Figure 8: Temporal correlation of the frequency of resistance with **community** use of antibiotics.

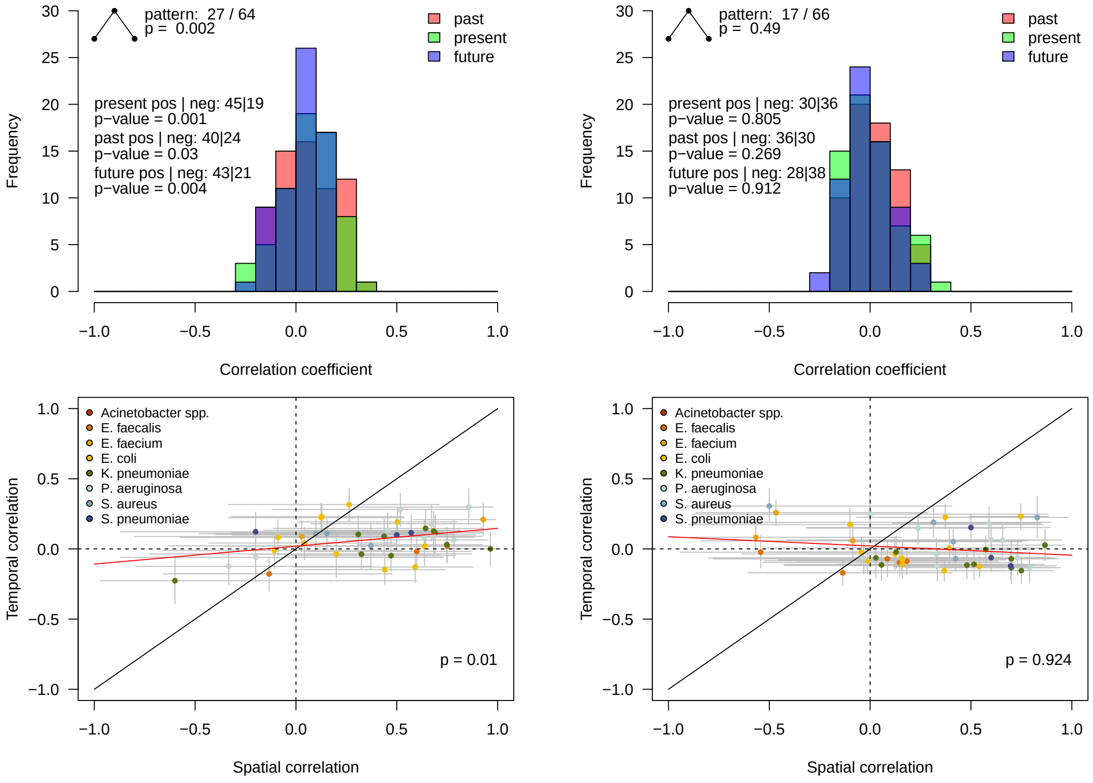


Figure 9: **Temporal correlation of the frequency of resistance with hospital antibiotic use (left) or community antibiotic use (right) across bug-drug combinations**. The top panels shows the distribution of the correlation coefficient of resistance with present antibiotic use, and antibiotic use shifted by $-1$ year in the past, and $+1$ year in the future. We also show the counts of combinations where temporal correlation is maximal in the present and associated p-value, and the counts of combinations with positive or negative temporal correlation (in the present, past and future) and associated p-value. A correlation maximal in the present indicate that the population is maximally adapted to its contemporaneous conditions. The frequent occurrence of this pattern further signals adaptation to fluctuating antibiotic use. The bottom panels show temporal correlation vs. spatial correlation across bug-drug combinations. A positive correlation between temporal and spatial correlations indicates that bug-drug combinations with more spatial adaptation also exhibit more temporal adaptation. The red line is a linear regression between these two quantities, with the p-value indicated. The two indicators suggest antibiotic resistance follows temporal fluctuations in the use of antibiotics in the hospital sector, but not in the community

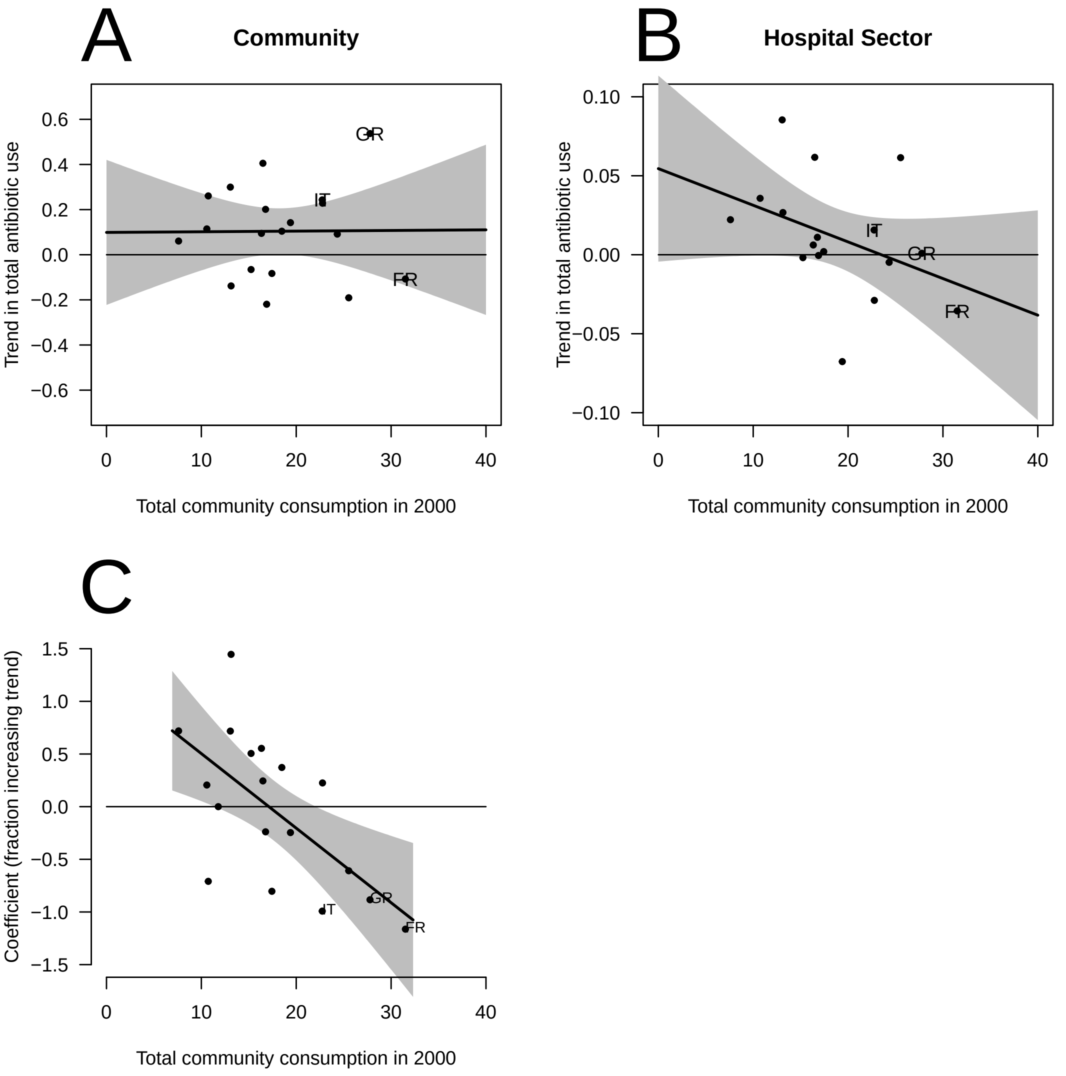


Figure 10: Trends in total antibiotic consumption (A, B) and in resistance (C) across countries, as a function of total community antibiotic use in 2000 (selecting any year in 1999-2003 leads to similar patterns). Trends in use are shown both for the community (A) and the hospital sector (B). Total use was calculated using the consumption of the main classes of antibiotics: tetracyclines, penicillins, other beta lactams, macrolides, quinolones. Points show the data, lines and shaded 95% confidence intervals are results from a linear regression. Antibiotic consumption tended to increase less in countries where it was high in 2000 in the hospital sector only (B). Countries with high antibiotic use in the community counted less increasing resistance trajectories (C). FR, IT, GR denote France, Italy, Greece.
